# Measurement of changes to the menstrual cycle: A transdisciplinary systematic review evaluating measure quality and utility for clinical trials

**DOI:** 10.1101/2024.04.04.24305348

**Authors:** Amelia C.L. Mackenzie, Stephanie Chung, Emily Hoppes, Alexandria K. Mickler, Alice F. Cartwright

## Abstract

Despite the importance of menstruation and the menstrual cycle to health, human rights, and sociocultural and economic wellbeing, the study of menstrual health suffers from a lack of funding, and research remains fractured across many disciplines. We sought to systematically review validated approaches to measure four aspects of changes to the menstrual cycle—bleeding, blood, pain, and perceptions—caused by any source and used within any field. We then evaluated the measure quality and utility for clinical trials of the identified instruments. We searched MEDLINE, Embase, and four instrument databases and included peer-reviewed articles published between 2006 and 2023 that reported on the development or validation of instruments assessing menstrual changes using quantitative or mixed-methods methodology. From a total of 8,490 articles, 8,316 were excluded, yielding 174 articles reporting on 94 instruments. Almost half of articles were from the United States or United Kingdom and over half of instruments were only in English, Spanish, French, or Portuguese. Most instruments measured bleeding parameters, uterine pain, or perceptions, but few assessed characteristics of blood. Nearly 60% of instruments were developed for populations with menstrual or gynecologic disorders or symptoms. Most instruments had fair or good measure quality or clinical trial utility; however, most instruments lacked evidence on responsiveness, question sensitivity and/or transferability, and only three instruments had good scores of both quality and utility. Although we took a novel, transdisciplinary approach, our systematic review found important gaps in the literature and instrument landscape, pointing towards a need to examine the menstrual cycle in a more comprehensive, inclusive, and standardized way. Our findings can inform the development of new or modified instruments, which—if used across the many fields that study menstrual health and within clinical trials—can contribute to a more systemic and holistic understanding of menstruation and the menstrual cycle.

## INTRODUCTION

### Menstrual health across disciplines

Menstruation and the wider menstrual cycle play a notable role in the health, human rights, and sociocultural and economic wellbeing of people who menstruate [1]. In addition, although its significance should not be utilitarianly reduced to only reproductive function, continuity of the human species would not occur without the menstrual cycle. Despite its importance, the study of menstruation and the menstrual cycle continues to suffer from a historical lack of funding and research across disciplines, including within the biological, clinical, public health, and social sciences. Within biomedical research, for example, a publication reporting on a recent technical meeting on menstruation convened by the United States (US) National Institutes of Health (NIH) decried a “lack of understanding of basic uterine and menstrual physiology” among researchers [2]. Indeed, many foundational, field-defining works have only recently emerged in the past five to ten years following increased attention to menstrual health, which the Global Menstrual Collective defined in 2021 as “a state of complete physical, mental, and social well-being and not merely the absence of disease or infirmity, in relation to the menstrual cycle” [3]. The contemporary growth of the menstrual health field is—at least partly—due to grassroots menstrual activism, which resulted in 2015 being labeled as “the year of the period” in the lay press [4]. Other examples of recent fundamental work within menstrual health across disciplines include recommendations for the menstrual cycle to be considered a vital sign and the advent of the field of critical menstruation studies [5,6]. Despite these recent efforts, insufficient research on menstrual health persists. In addition, the study of menstrual health remains fractured across many fields and disciplines, many of which are siloed despite adjacent or even overlapping subject matters (e.g., menstrual health and hygiene within wider sexual and reproductive health; or gynecology, endocrinology, and many other specialties within medicine) [7,8]. As a result, we still lack a complete, systemic, and holistic understanding of menstruation and the wider menstrual cycle.

The type of interdisciplinary, comprehensive global efforts needed to address such large gaps in menstrual health research can greatly benefit from standardization—of terminology, of measurement, of analysis, and of outcomes or indicators. The widest global effort at standardization to date has taken place within medicine; the International Federation of Gynecology and Obstetrics (FIGO) established clinical standards of normal and abnormal uterine bleeding occurring outside of pregnancy via a consensus-building process over a series of years [9–12]. These FIGO standards dictate four parameters for menstrual bleeding: the frequency, duration, volume, and regularity of bleeding. FIGO defines normal uterine bleeding as bleeding occurring every 24-38 days (frequency), bleeding lasting no more than 8 days (duration), bleeding of a ‘normal’ amount as defined by the patient that does not interfere “with physical, social, emotional, and/or material quality of life” (volume), and bleeding within a menstrual cycle that only varies in length by plus or minus 4 days (regularity). FIGO further defines bleeding outside these normal parameters as abnormal uterine bleeding, which is divided into standard categories based on whether it is acute or chronic and the source or etiology of the abnormality according to the acronym PALM-COEIN (i.e., <u>P</u>olyp, <u>A</u>denomyosis, <u>L</u>eiomyoma, <u>M</u>alignancy and hyperplasia, <u>C</u>oagulopathy, <u>O</u>vulatory dysfunction, <u>E</u>ndometrial disorders, Iatrogenic, and <u>N</u>ot otherwise classified). Other examples of efforts at standardization include menstrual hygiene indicators within the Water, Sanitation and Hygiene (WASH) field and defining how contraception can impact the menstrual cycle and analyzing these data in contraceptive studies [13–18].

Related to terminology, this review uses the phrase, “people who menstruate”, which we define as those who can menstruate, do menstruate, or have menstruated. Although people who menstruate may or may not identify as women or girls, and not all women and girls menstruate [19], we do use the terms ‘women’ and ‘girls’ in some instances, especially when citing primary literature and because menstrual health cannot “be adequately addressed without attention to the gender norms and dynamics experienced by individuals in the cultures and communities in which they live” [7]. As much as possible, however, we use gender inclusive terms and other people-first language.

### Review scope

To aid in efforts for standardized measurement across the study of menstruation and the menstrual cycle, we systematically reviewed approaches to measure four aspects of changes to the menstrual cycle: bleeding, blood, pain, and perceptions of bleeding, blood, or pain. We use the term ‘**menstrual changes**’ to refer to these four aspects for the remainder of the paper. We sought to include all types of measures or methods for assessing menstrual changes (e.g., quantitative assays, biomarkers, data reported by clinicians, researchers, or directly by the person who menstruates). We use the term ‘**instruments**’ to refer to any of these measures or methods for the remainder of the paper. Our aim was to identify any instruments that have been developed and validated within any field of study to measure menstrual changes, examine how these instruments measured menstrual changes, and assess the measure quality of the identified instruments and their utility for the clinical trial context.

Related reviews have been conducted: (a) within fields such as menstrual hygiene or the study of heavy menstrual bleeding (HMB) [20,21]; (b) to measure single parameters like volume of menstrual blood loss 22]; and (c) for specific approaches like pictorial methods to diagnose HMB [23]. However, given the gaps and silos within menstrual health research, our aim was to conduct an expansive and transdisciplinary review to inform more standardized measurement across menstrual health research and clinical trials. For this reason, we sought to include menstrual changes caused by any etiology or source. There are many factors that can result in menstrual changes, including those endogenous and exogenous to the person who menstruates. Examples of these etiologies or sources include menstrual or gynecologic disorders like adenomyosis, use of hormonal or intrauterine contraceptives, use of other drugs or devices to treat or prevent disease, environmental exposures, infectious disease, injury, coagulation disorders, and diet and exercise. We are not aware of any previous efforts to look at menstrual changes across disciplines in this way.

### Clinical trial context

As mentioned, one area for which we intend our review to be quite relevant is for data collection in clinical trials, although our broad approach does not preclude the use of our results to inform the measurement of menstrual changes across other research contexts. The importance of data on menstrual changes in the clinical trial context was recently highlighted during the introduction of COVID vaccinations. Because vaccine trials did not collect data on the impact to the menstrual cycle or menopausal uterine bleeding, there were concerns among vaccinated people who menstruate when they experienced these changes, which can erode trust in clinical research and public health interventions [24–28]. As authors working across various sexual and reproductive health spaces, our interest in conducting this review stemmed from a shared goal to improve and standardize the measurement of menstrual changes in contraceptive clinical trials; however, our broad methodological approach permits the utility of our findings across all clinical trials.

Clinical trials, and the preclinical research that precedes them, collect data on key organ functioning and vital signs as part of standard toxicology and pharmacodynamics. Given the importance of the menstrual cycle, it may seem surprising that data on how investigational drugs may impact the menstrual cycle are not already routinely collected in clinical trials; however, research typically reflect the people, priorities, and purposes of those within the clinical trial ecosystem—that is, the individuals and systems that fund clinical research, conduct clinical trials, and regulate the drugs tested in trials, as well as the individuals who participate in trials. Historically, there has been an underrepresentation of people who menstruate within the clinical trial ecosystem [29]. This exclusion is true for much of the preclinical research that informs clinical trials across many biomedical fields as well, and even cell lines used in *in vitro* studies are predominantly derived from male animals [30,31]. Although proof-of-concept studies for drugs intended for use in women that are known to impact the menstrual cycle, such as hormonal contraceptives, do typically use female animals when the model organism has an estrous or menstrual cycle, other preclinical research disproportionately relies on only male animals. Using both female and male animals in the research that informs clinical trials, however, could provide early indications of any impacts on cycles, as well as many other sex-specific effects or differences. Despite decades of concrete efforts, sex and gender disparities persist in the clinical trial ecosystem [32–34].

Within the current clinical trial context, another element relevant to our review is how trials typically incorporate outcomes, like menstrual changes, that are reported by trial participants. The US Food and Drug Administration (FDA) and NIH refer to these data as patient-reported outcomes (PROs), which they define as “a measurement based on a report that comes directly from the patient (i.e., study subject) about the status of a patient’s health condition without amendment or interpretation of the patient’s response by a clinician or anyone else.” PROs can include “symptoms or other unobservable concepts known only to the patient (e.g., pain severity or nausea) [that] can only be measured by PRO measures,” as well as “the patient perspective on functioning or activities that may also be observable by others” [35]. Unless an assay or biomarker are used, all outcomes on menstrual changes are reported by the person who menstruates and, therefore, are PROs. The FDA has a series of methodological guidance documents on the development, validation, and use of PROs in clinical trials as part of patient-focused drug development efforts [36–39].

### Review questions and objective

Given the aim of the review, our review questions were: (a) What instruments have been developed to assess menstrual changes caused by any etiology or source? and (b) What is the quality of these instruments and their utility for clinical trials? The objective of our systematic review was to compile a complete list of validated instruments used to measure menstrual changes with an assessment of their quality and clinical trial utility.

## MATERIALS AND METHODS

We conducted our systematic review in alignment with Preferred Reporting Items for Systematic Reviews and Meta-Analysis (PRISMA) guidelines [40–42], including a protocol registered in PROSPERO (Protocol ID: CRD42023420358) [43]. A completed PRISMA checklist for this review is in Table S1, and Appendix S2 includes additional details on the search strategy, inclusion/exclusion criteria, title/abstract screening, full text review, data extraction, and data analysis.

### Search strategy

We searched for peer reviewed articles in the MEDLINE and Embase literature databases and for any relevant instruments measuring menstrual changes in four instrument databases: (a) the NIH Common Data Element (CDE) Repository [44], (b) the COSMIN database of systematic reviews of outcome measurement instruments [45], (c) the Core Outcome Measures in Effectiveness Trials (COMET) Database [46], and (c) ePROVIDE databases [47]. Table 1 shows the final search strategy for MEDLINE, and Appendix S2 includes search strategies for other databases. We uploaded articles from the literature database searches and articles for any relevant instruments identified via the instrument databases into Covidence [48]. Following screening and review of these articles in Covidence, we identified relevant review articles and extracted primary articles published since 1980 from those reviews. During data extraction, we identified any original articles for instruments developed before 2006. We uploaded these primary articles and original development articles into Covidence for screening.

**Table 1.**
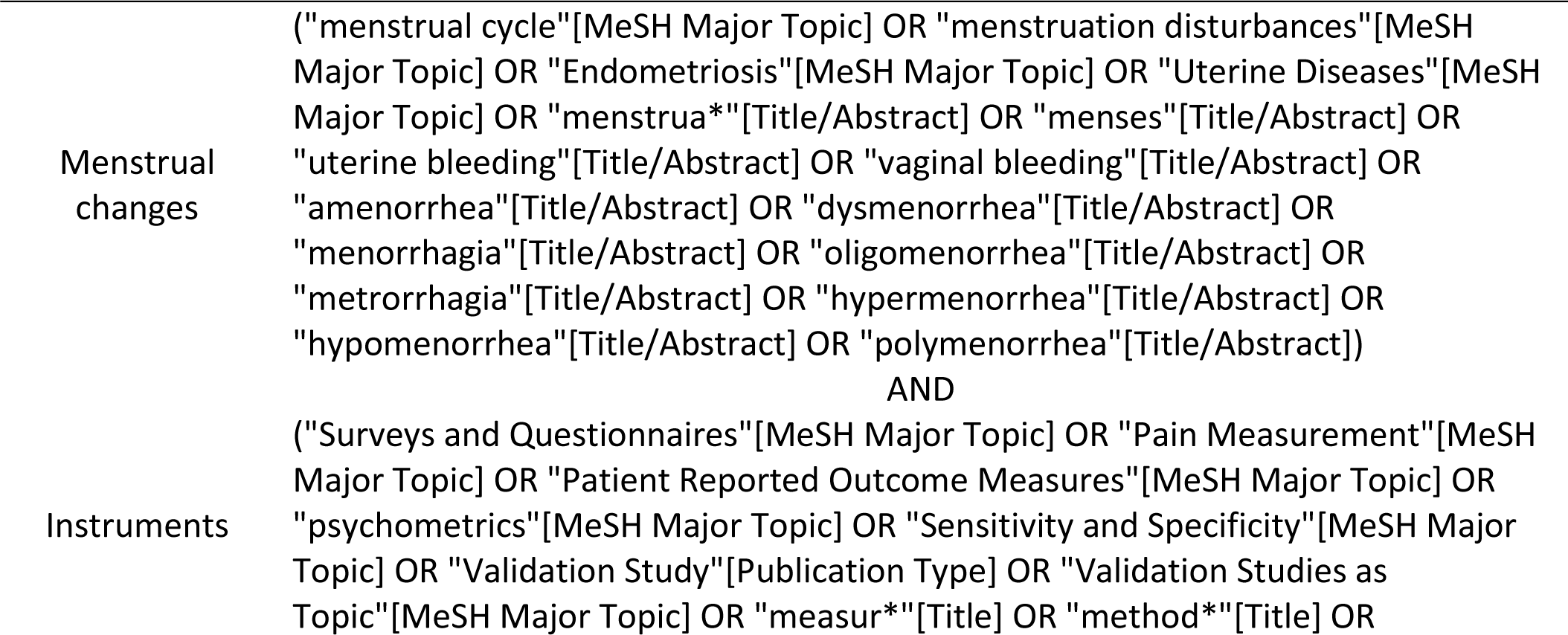
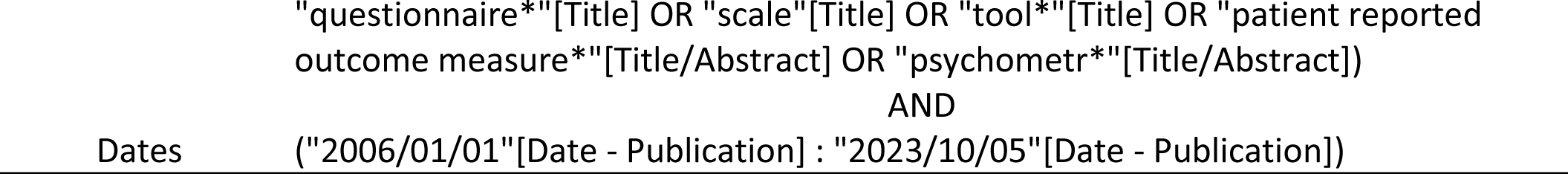
MEDLINE search strategy.

### Inclusion/exclusion criteria

We included all peer-reviewed articles—including those with prospective, retrospective, or cross-sectional study designs, and review papers—that met our inclusion and did not meet our exclusion criteria, listed in Table 2.

**Table 2.**
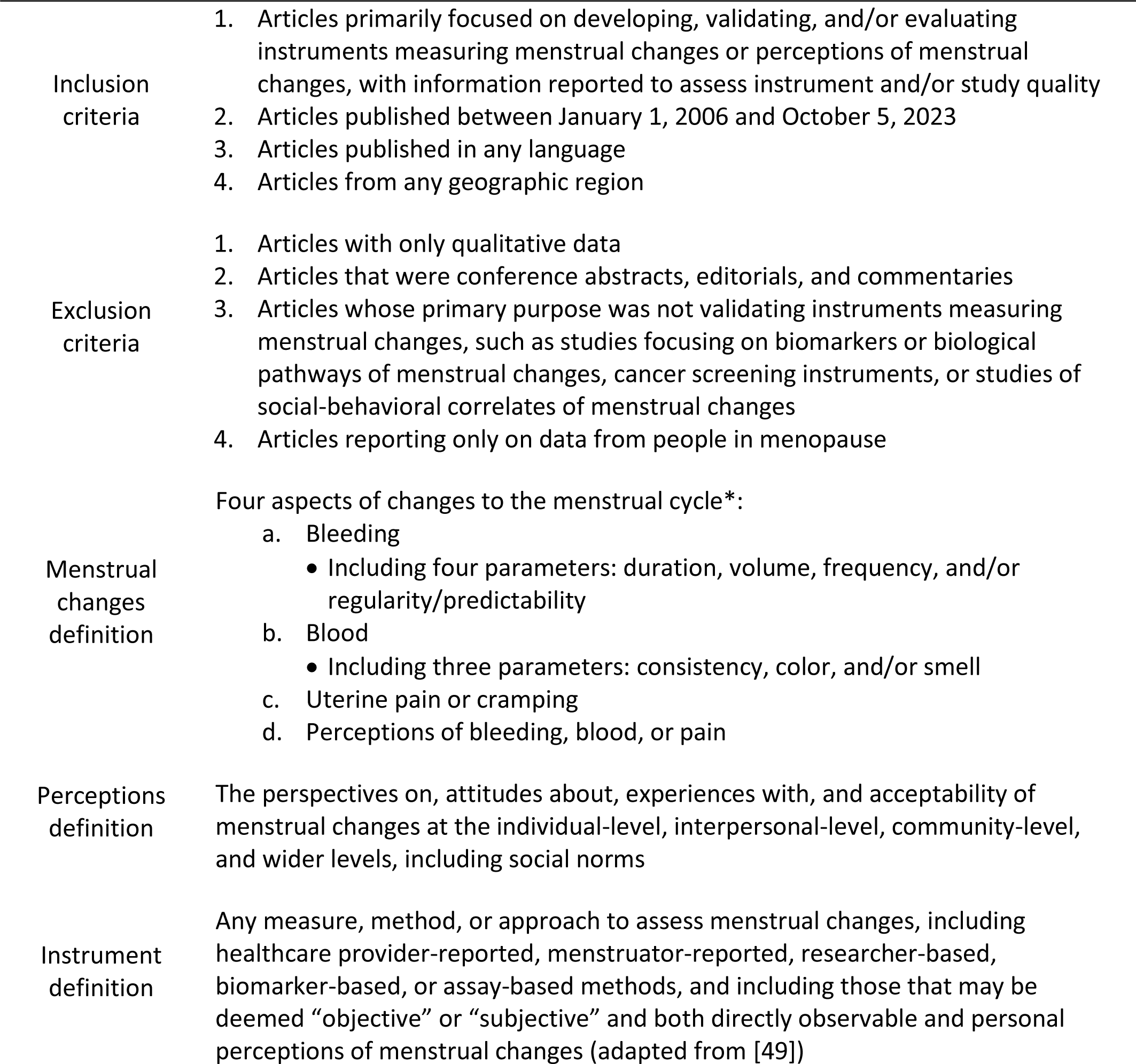
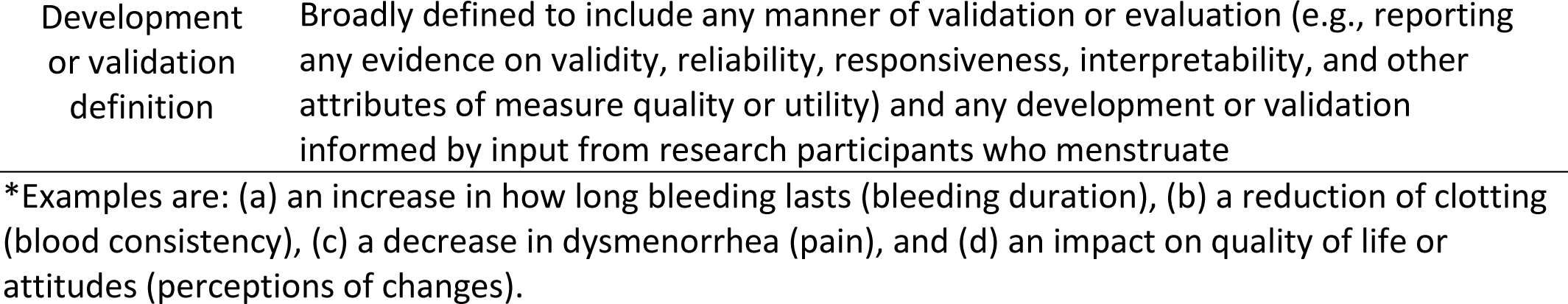
Inclusion and exclusion criteria and related definitions.

|  |  |
| --- | --- |
| Inclusion criteria | Articles primarily focused on developing, validating, and/or evaluating instruments measuring menstrual changes or perceptions of menstrual changes, with information reported to assess instrument and/or study quality<br>Articles published between January 1, 2006 and October 5, 2023<br>Articles published in any language<br>Articles from any geographic region |
| Exclusion criteria | Articles with only qualitative data<br>Articles that were conference abstracts, editorials, and commentaries<br>Articles whose primary purpose was not validating instruments measuring menstrual changes, such as studies focusing on biomarkers or biological pathways of menstrual changes, cancer screening instruments, or studies of social-behavioral correlates of menstrual changes<br>Articles reporting only on data from people in menopause |
| Menstrual changes definition | Four aspects of changes to the menstrual cycle*:<br>Bleeding<br>Including four parameters: duration, volume, frequency, and/or regularity/predictability<br>Blood<br>Including three parameters: consistency, color, and/or smell<br>Uterine pain or cramping<br>Perceptions of bleeding, blood, or pain |
| Perceptions definition | The perspectives on, attitudes about, experiences with, and acceptability of menstrual changes at the individual-level, interpersonal-level, community-level, and wider levels, including social norms |
| Instrument definition | Any measure, method, or approach to assess menstrual changes, including healthcare provider-reported, menstruator-reported, researcher-based, biomarker-based, or assay-based methods, and including those that may be deemed “objective” or “subjective” and both directly observable and personal perceptions of menstrual changes (adapted from [49]) |
| Development or validation definition | Broadly defined to include any manner of validation or evaluation (e.g., reporting any evidence on validity, reliability, responsiveness, interpretability, and other attributes of measure quality or utility) and any development or validation informed by input from research participants who menstruate |

\*Examples are: (a) an increase in how long bleeding lasts (bleeding duration), (b) a reduction of clotting (blood consistency), (c) a decrease in dysmenorrhea (pain), and (d) an impact on quality of life or attitudes (perceptions of changes).

### Title/abstract screening, full text review, and data extraction

We held weekly author meetings to discuss progress, questions, and discordance, and to document decisions in a shared Word document. We began title/abstract screening with an ‘inter-reviewer reliability’ meeting where all authors completed title/abstract screening on the same 50 articles to establish and confirm group standards. Then, two authors independently screened each remaining title/abstract and two authors independently reviewed each relevant full text in Covidence. We resolved any discordance during weekly meetings via consensus conversations. We conducted data extraction in Excel using a template data extraction form that collected information on article information, study design, sample information, details on the instrument, measure quality attributes, and clinical trial utility attributes. For articles not in English, we used the text translation feature of Google Translate to review titles and abstracts, we used the document translation feature of Google Translate and/or consulted a fluent colleague to review full text articles, and we completed data extraction with a fluent colleague for included articles.

For assessing measure quality and clinical trial utility, we followed the recent Patient-Reported Outcomes Tools: Engaging Users and Stakeholders (PROTEUS) Consortium recommendations to use the International Society for Quality of Life Research (ISOQOL) standards for PRO measures [50,51]. We made two adjustments to the ISOQOL standards: (a) we added an attribute on sensitivity of questions given the topic of menstruation has a noted amount of stigma surrounding it [52]; and (b) we separated out participant burden from investigator burden given these two can differ greatly for instruments measuring menstrual changes. We categorized six attributes as related primarily to the quality of the instrument (i.e., **measure quality**: conceptual/measurement model, reliability, content validity, construct validity, responsiveness, and sensitive nature of questions) and four attributes as related primarily to the utility of the instrument in clinical trials (i.e., **clinical trial utility**: interpretability of results, the transferability of the instrument, participant burden, and investigator burden). We scored each attribute of measure quality and clinical trial utility on a scale from 0 to 3 (0= **no data,** 1=**poor**, 2=**fair**, and 3=**good**) based on criteria in line with ISOQOL standards [51] that were reviewed by measurement and clinical experts at FHI 360 and within a related global task force. We show the measure quality and clinical trial utility attributes and scoring criteria in Table 3, and Appendix S2 includes details on the other fields of the data extraction form.

**Table 3.**
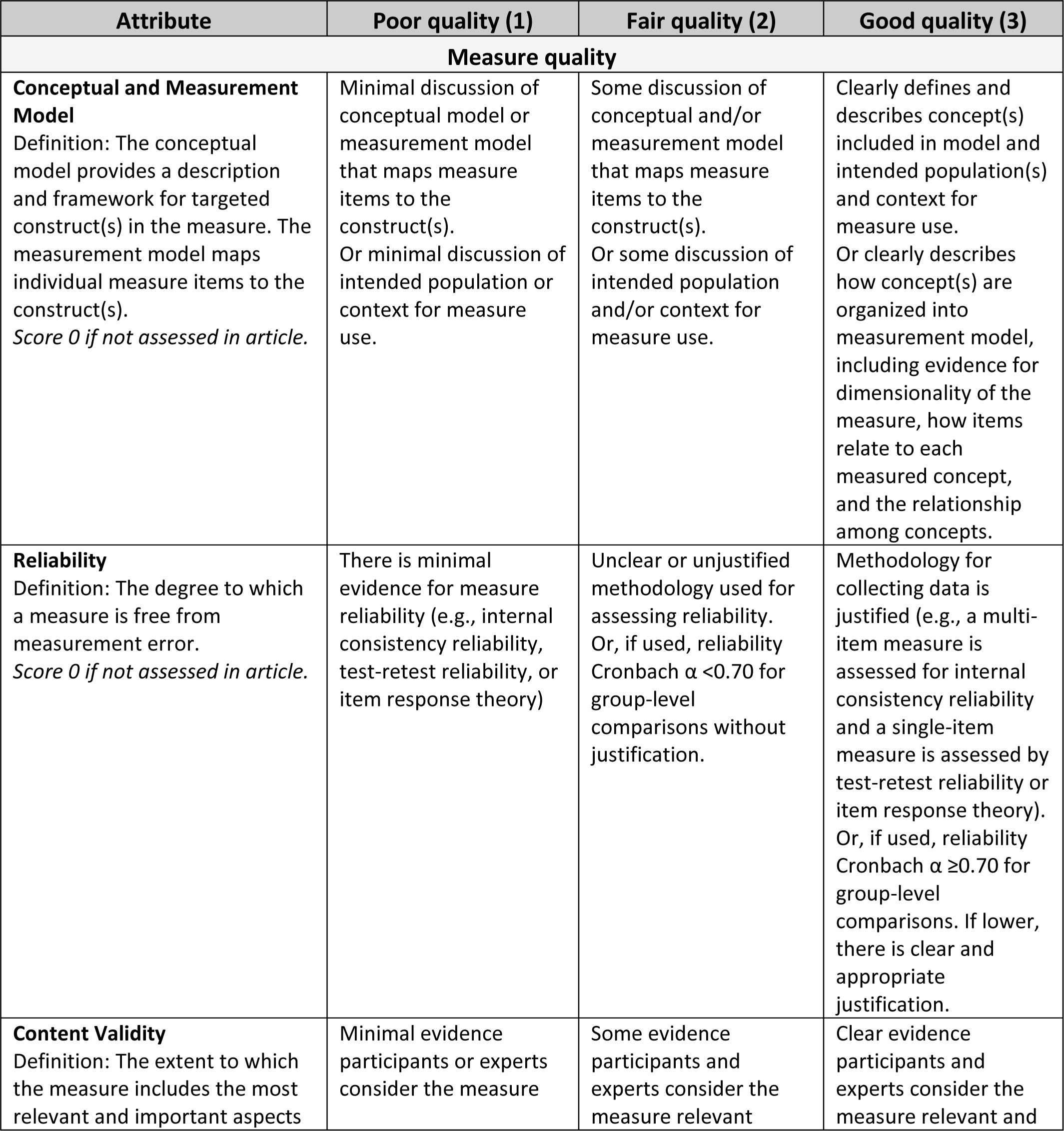
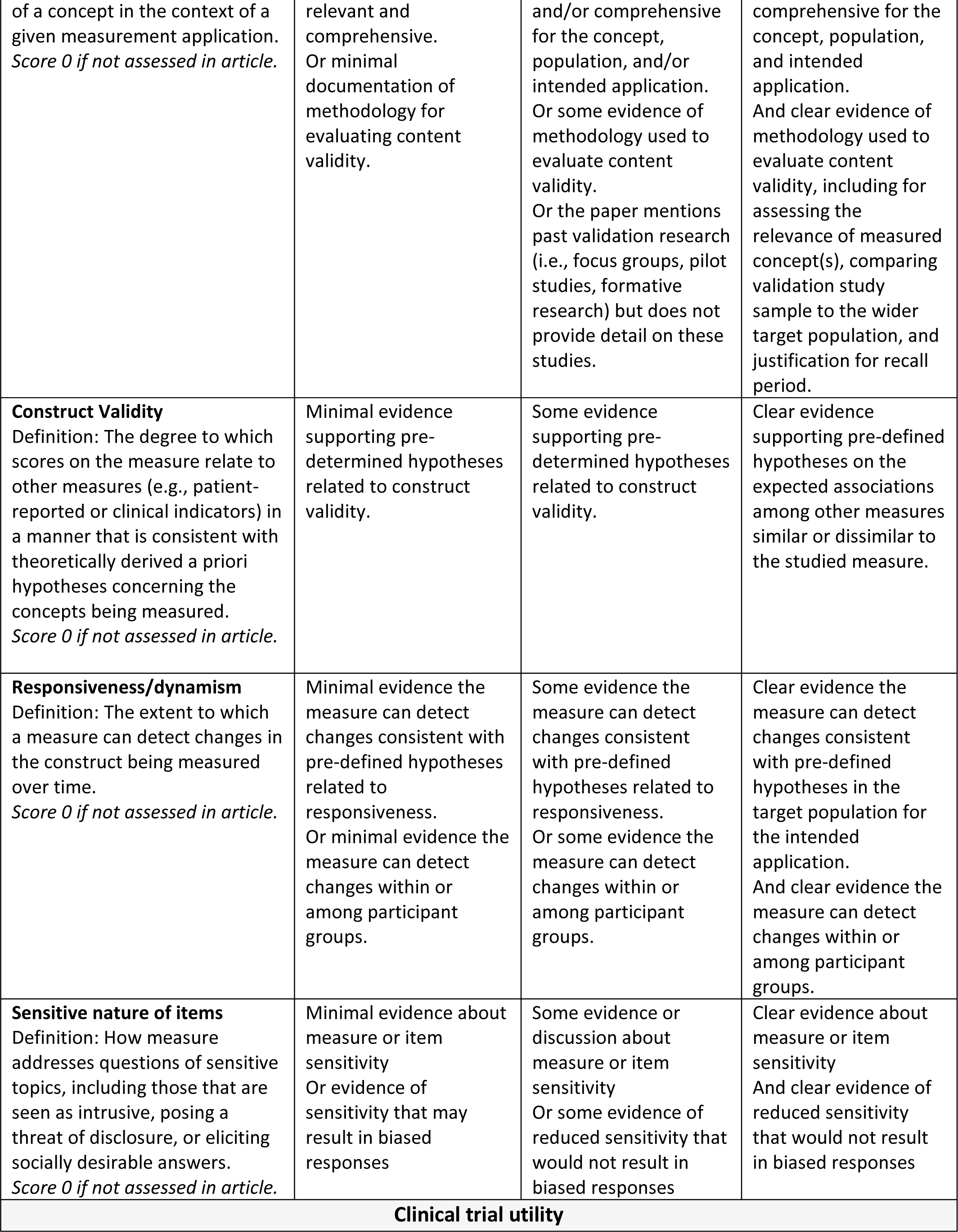
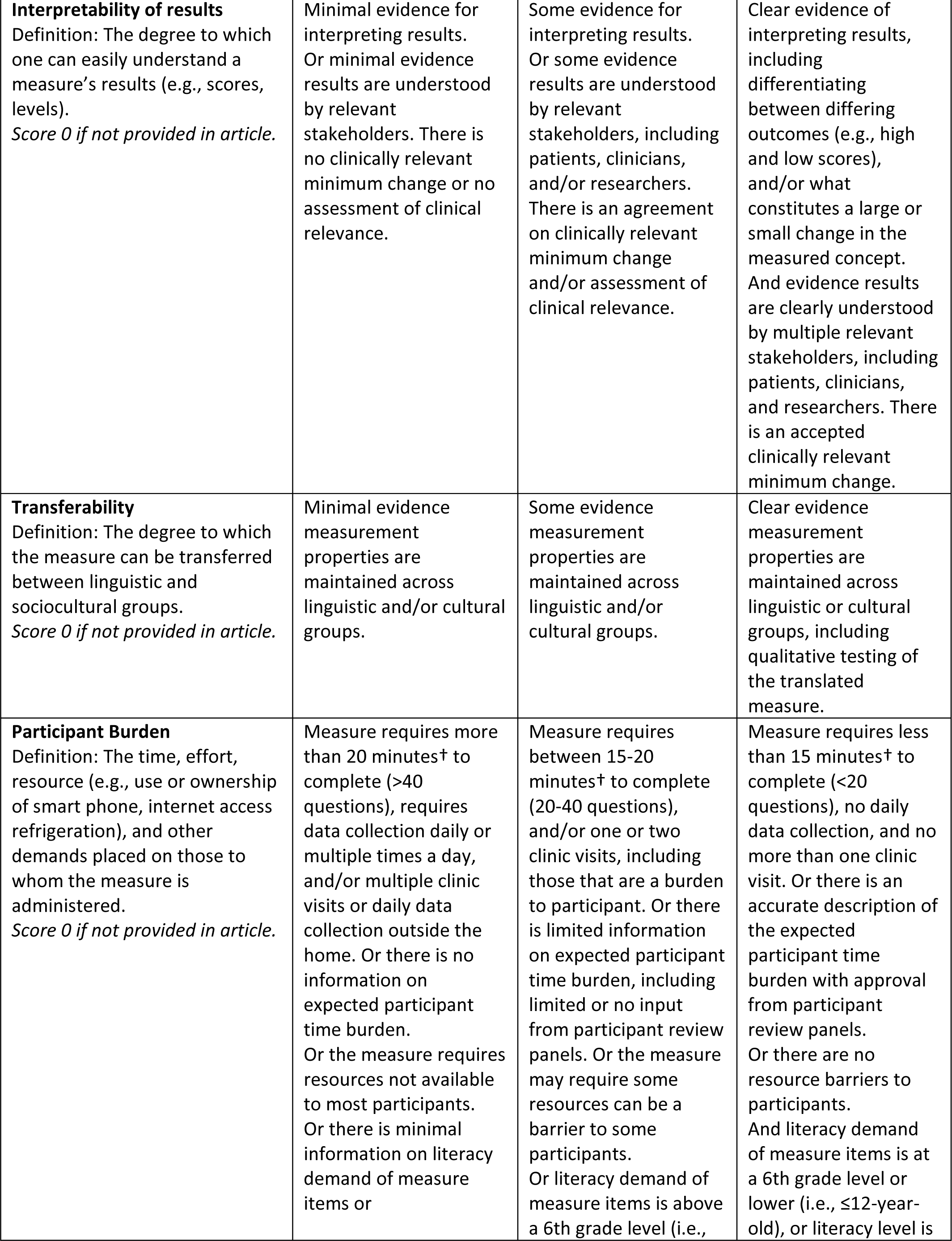
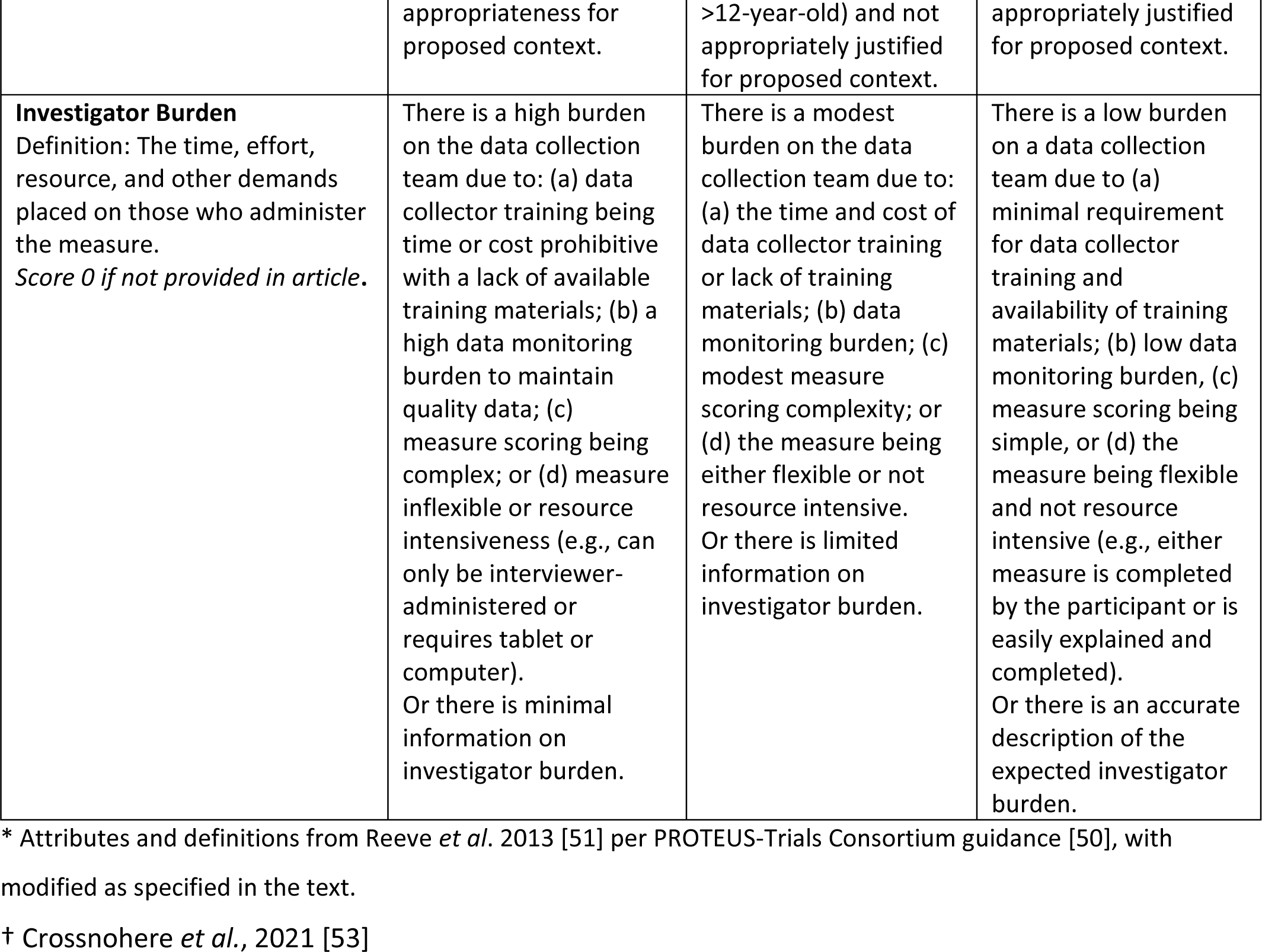
Measure quality and clinical trial utility scoring criteria*.

### Data analysis

We conducted data analysis in Excel and included counts and frequencies, as well as specific analyses to assess instrument measure quality and clinical trial utility. For the **measure quality score** and **clinical trial utility score** of an instrument, we used an average of the highest score for each attribute of measure quality or clinical trial utility across all articles on an instrument. Because instruments could have more than one article providing data on measure quality and/or clinical trial utility and not every article evaluated all attributes of an instrument, we did not include scores of zero (i.e., no data reported) in these averages. To reflect these differences in the number of articles and attributes reported in the article(s), we also calculated a total **evidence score**, which was the total of all scores—including zeros— across all attributes of measure quality and clinical trial utility. The total evidence scores, therefore, ‘penalize’ instruments for a lower level of evidence due to fewer articles or less attribute data and vice versa.

These three scores—measure quality (ranging from 1-3), clinical trial utility (ranging from 1-3), and total evidence (ranging 0+)—reflect different dimensions of an instrument. For example, two instruments might both have a score of 2.5 for measure quality, but one instrument might have an evidence score of 10 and the other, 100, indicating the latter has considerably more evidence and likely more certainty in the measure quality score. Alternately, two instruments may have similar measure quality and evidence scores, but one may have a clinical trial utility score of 1 and the other a score of 3, indicating the latter is likely better suited for use in clinical trials despite the similar levels of measure quality and evidence.

## RESULTS

### Search results

Across databases, our searches yielded 8,490 articles. We removed 376 duplicates, excluded 7,704 articles during title and abstract screening, and excluded 236 articles during full text review. In total, we identified 174 relevant full text articles. We present additional details on our search results and screening in the PRISMA diagram in Figure 1.

**Fig. 1.**
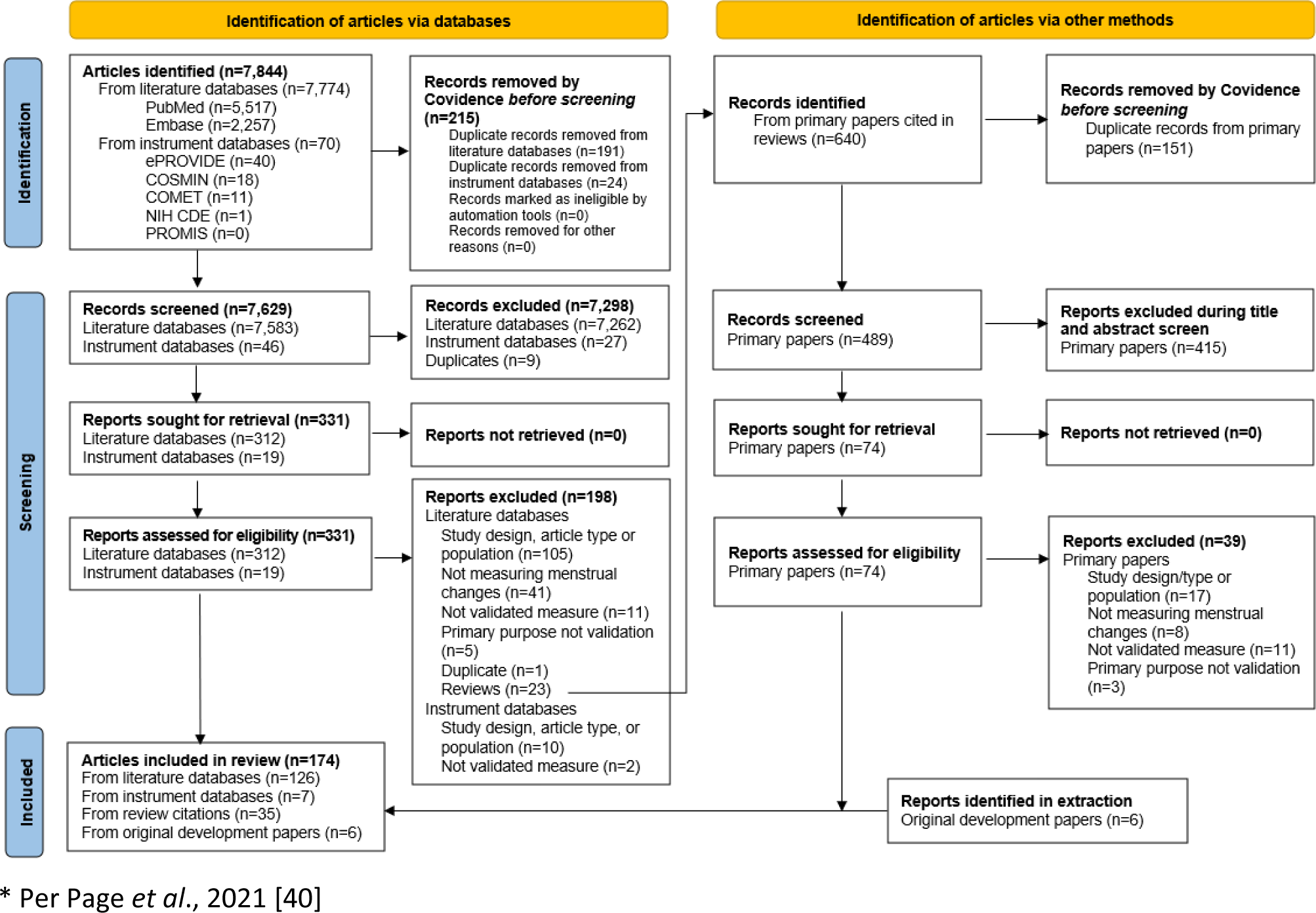
PRISMA Diagram*.

We found some similarities across papers that we excluded for not meeting our inclusion criteria. For example, we excluded conference presentations that never became full papers, studies that focused on validating instruments among only menopausal populations (e.g., [54,55]), and studies that only validated surgical or treatment outcomes (e.g., [56,57]). In addition, there were two recent papers on core outcome sets for HMB and endometriosis relevant to the wider topic of measuring changes to the menstrual cycle, but we excluded them because there were no instrument details to extract [58,59].

### Included article characteristics

Over 85% of the 174 articles were from either Europe (43%), North America (32%) or Asia (13%), and there were less than 15 articles from South America (n=13), from the Middle East (n=11), from Oceania (n=8) and from Africa (n=5). Just under half of articles were from only the United States (28%) or the United Kingdom (16%), although we did identify articles from a total of 50 countries. Nearly all articles were in English—even those reporting on instruments in other languages—except for two in Portuguese [60,61]. The most common study designs were cross-sectional or prospective cohort. We present details of all 174 included articles in Table S3.

### Instrument characteristics

From the 174 included articles, we extracted 94 instruments. Almost three quarters (72%, n=68) were full instruments, collecting data on one or more menstrual change. Nearly a quarter (n=21) were broader instruments that included sub-scales (9%, n=8) or a small number of items (14%, n=13) on menstrual changes. Five percent (n=5) were general instruments validated in menstruating populations on one or more menstrual change. The instruments with the most articles in our review were the Endometriosis Health Profile-30 (EHP-30; 20 articles), the Pictorial Blood Loss Assessment Charts & Menstrual Pictograms (PBAC; 11 articles), the Uterine Fibroid Symptom and Quality of Life questionnaire (UFS-QOL; 9 articles), the Polycystic Ovary Syndrome Quality of Life scale (PCOS-QOL; 8 articles), and the Endometriosis Health Profile-5 (EHP-5), Menstrual Attitudes Questionnaire (MAQ), and menstrual collection (5 articles each). About a third (38%, n=26) of full instruments used electronic data collection, and almost all full instruments (97%, n=66) were completed by only the patient/participant who menstruated (i.e., they were PROs per the FDA and NIH definition). We present the list of the 68 full instruments and instrument characteristics in Table 4. The remainder of results reported below are for these full instruments, with details on the sub-scales, items, and general instruments in Table S4.

**Table 4:**
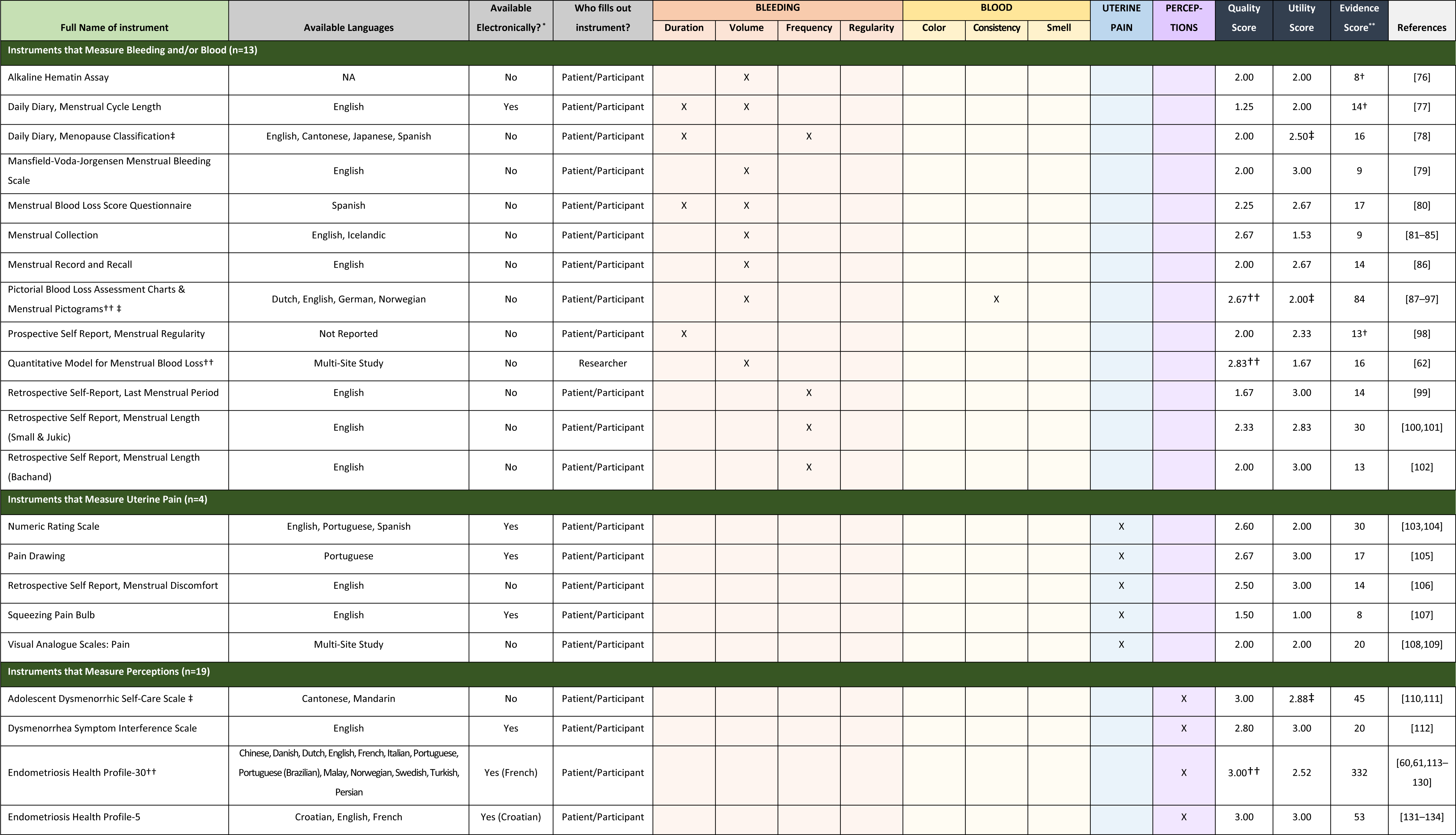
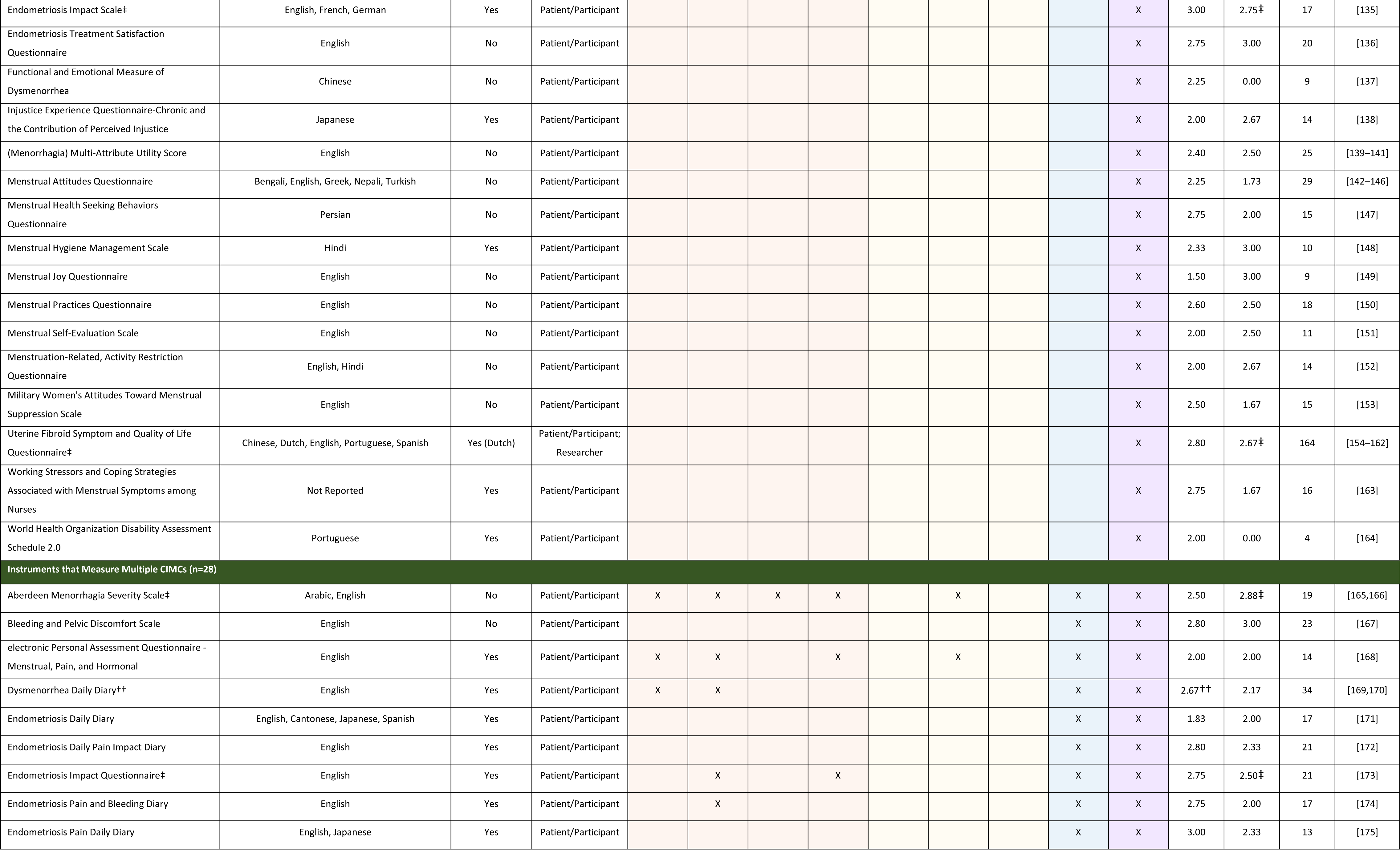
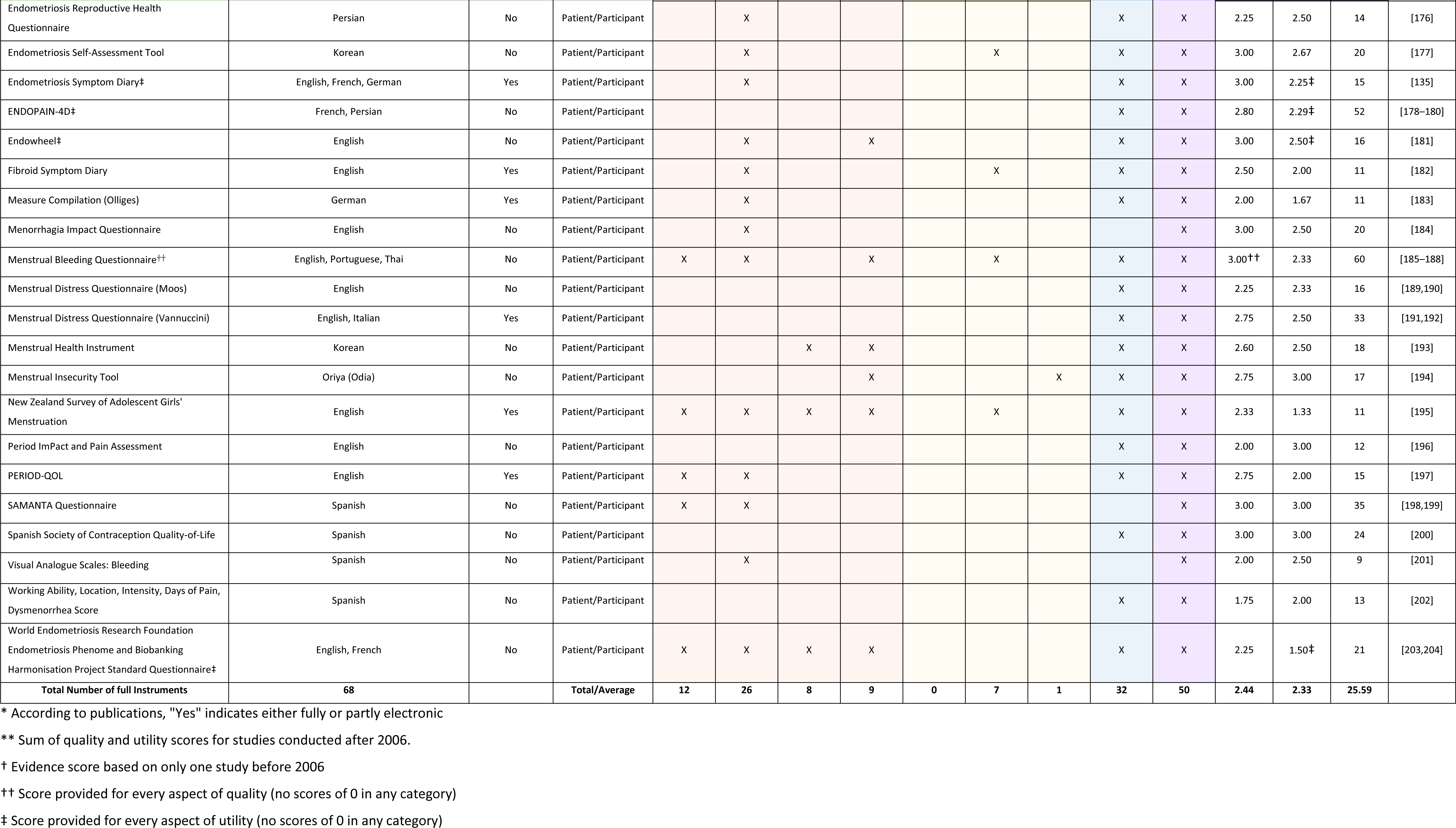
List of full instruments and characteristics.

### Language(s)

Of the 68 full instruments, two-thirds were in English (66%, n=45), followed by Spanish (13%, n=9), French (9%, n=6), and Portuguese (9%, n=6); however, we identified instruments in 28 languages. About forty percent of instruments (41%, n=28) were only in English, although about a quarter of instruments (26%) were in more than one language, and six instruments were in at least 4 languages. These instruments included the EHP-30 (13 languages), UFS-QOL (5 languages), MAQ (5 languages), PBAC (4 languages), Endometriosis Daily Diary (EDD; 4 languages), and the Daily Diary (4 languages).

### Specific Populations

Nearly 60% (n=40) of the 68 full instruments were developed and/or validated in populations with menstrual or gynecologic disorders or symptoms (i.e., 18 for endometriosis, 10 for HMB, 9 for dysmenorrhea, and 3 for uterine fibroids). Less than a quarter (24%, n=16) of full instruments were developed for and validated with adolescents (mean ages less than 18, n=10) or young people (mean ages early 20s, n=6). Three full instruments were specifically developed for those in perimenopause. A few instruments were developed or validated in populations of athletes or people in the military. No instruments or articles indicated inclusion of trans and gender nonbinary people who menstruate.

### Menstrual change(s) measured

Among the 68 full instruments, half (49%, n=33) measured bleeding, nearly half (47%, n=32) measured uterine cramping or pain, and almost three quarters (74%, n=50) measured perceptions. Only eight (12%) measured blood characteristics. Three instruments assessed all four of the parameters of bleeding—duration, volume, frequency, and regularity/predictability (i.e., the Aberdeen Menorrhagia Severity Scale [AMSS], the New Zealand Survey of Adolescent Girls’ Menstruation, and the World Endometriosis Research Foundation Endometriosis Phenome and Biobanking Harmonisation Project Standard Questionnaire [WERF EPHect EPQ-S]). No instrument assessed all three parameters of blood— color, consistency, and smell.

Across the four aspects of menstrual changes (i.e., bleeding, blood, pain, and perceptions), no instrument measured all parameters for each aspect, and only seven instruments measured at least a single parameter of each aspect. These instruments were the AMSS; electronic Personal Assessment Questionnaire - Menstrual, Pain, and Hormonal (ePAQ-MPH); Endometriosis Self-Assessment Tool (ESAT); Fibroid Symptom Diary (FSD); Menstrual Bleeding Questionnaire (MBQ); Menstrual Insecurity Tool; and the New Zealand Survey of Adolescent Girls’ Menstruation.

### How instruments measured menstrual changes

We present in Table 5 details on how the 68 full instruments measured bleeding (i.e., the four parameters of duration, volume, frequency, and regularity/predictability), blood (i.e., the three parameters of color, consistency, and smell), uterine cramping/pain, and perceptions.

**Table 5:**
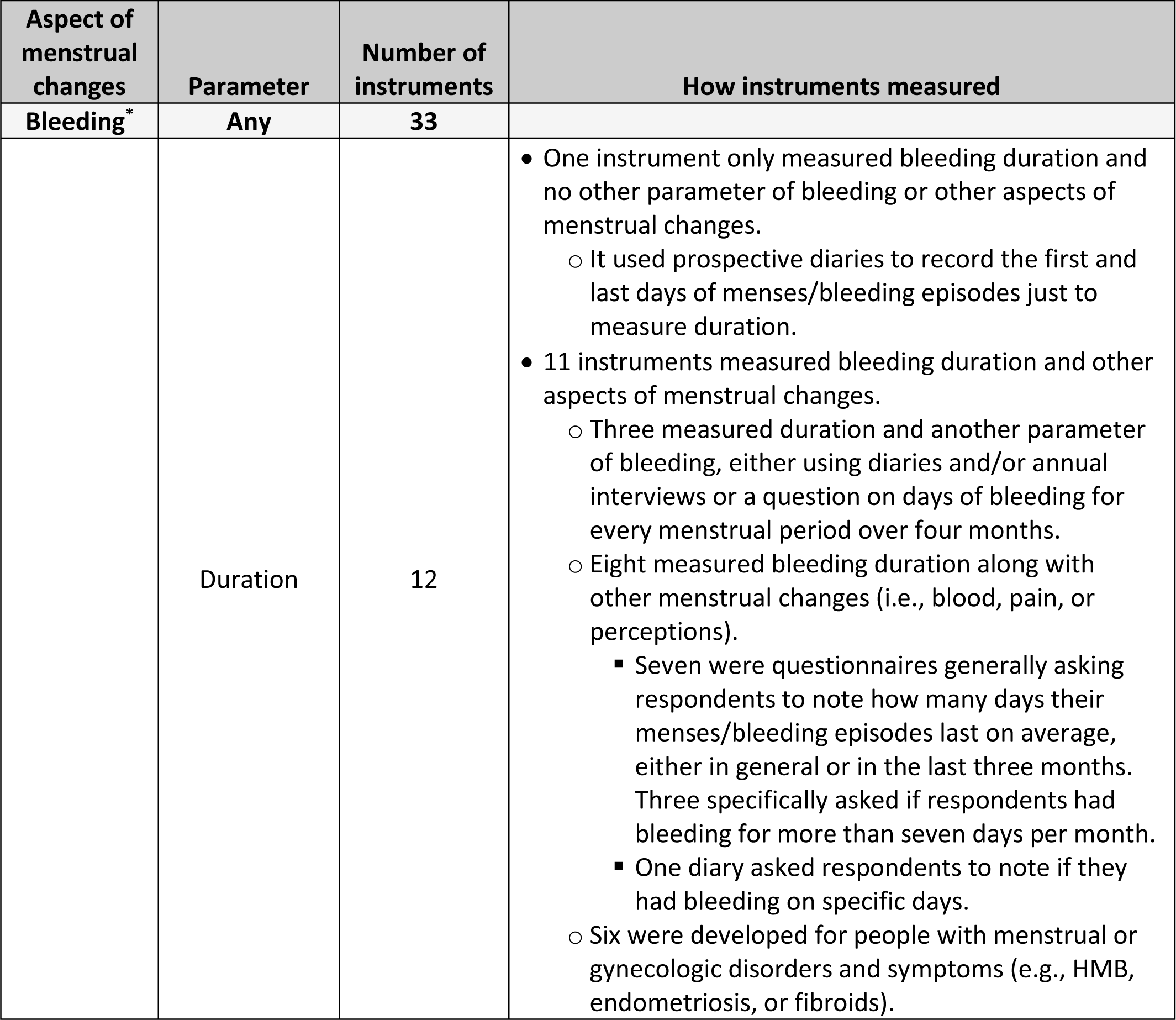
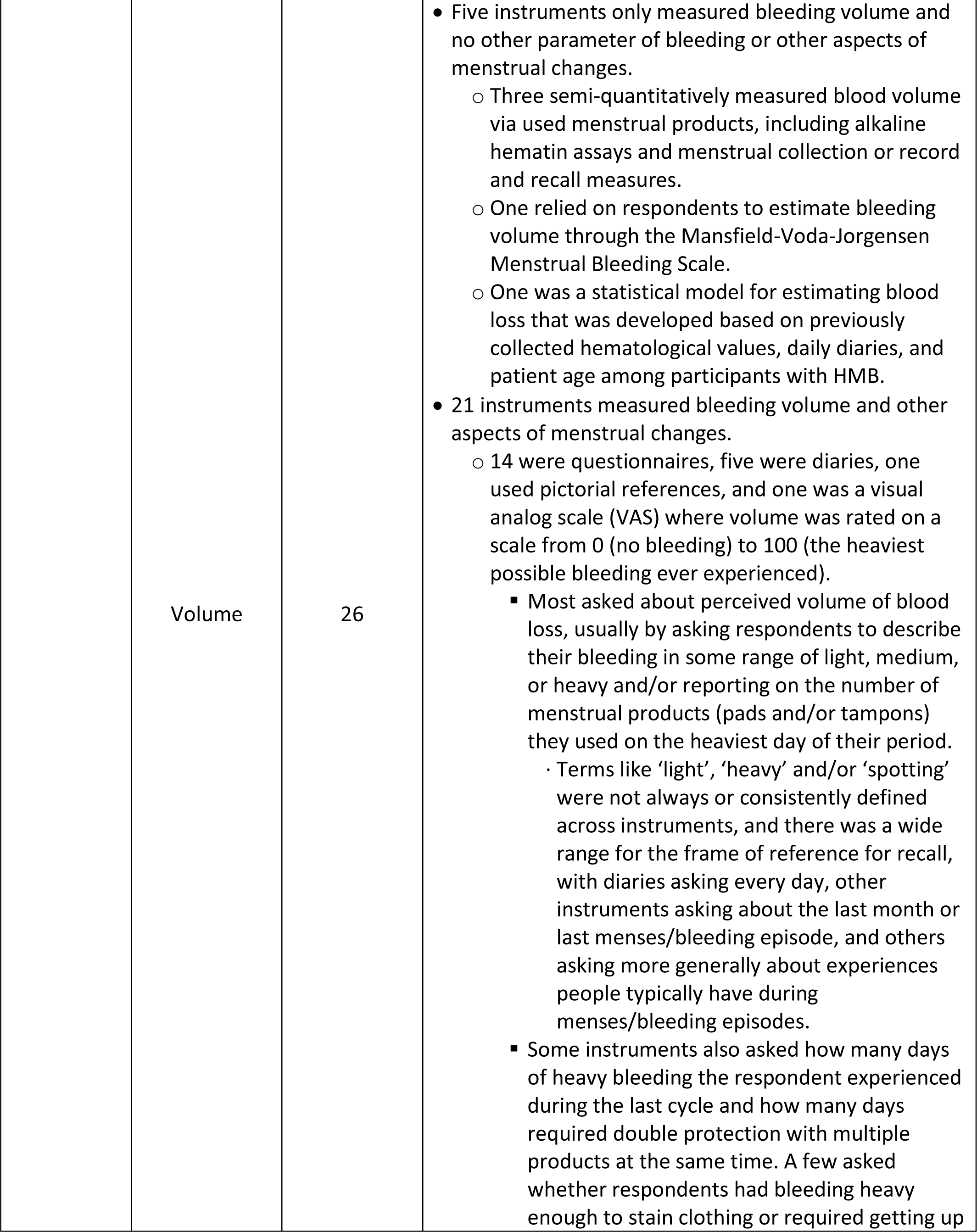
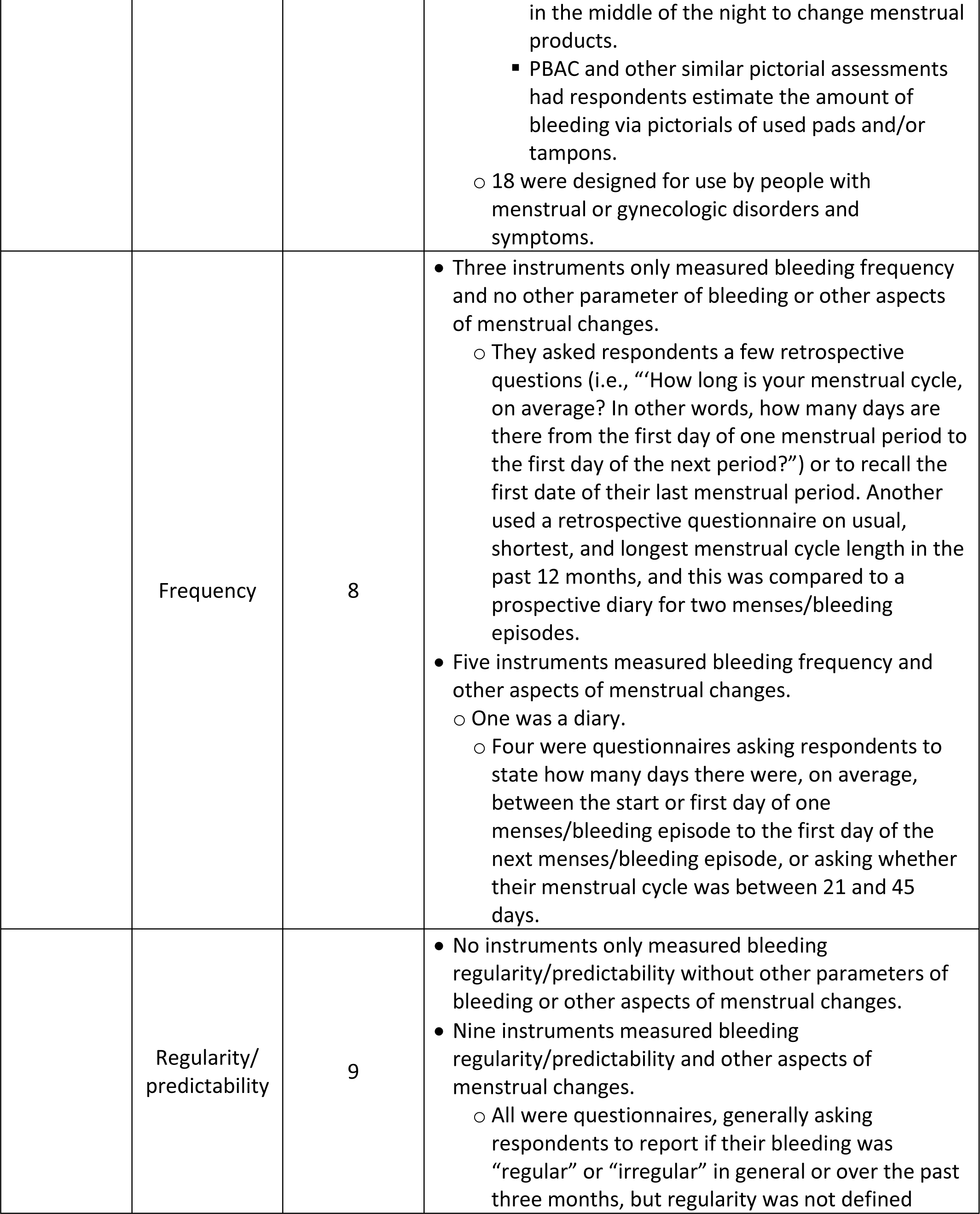
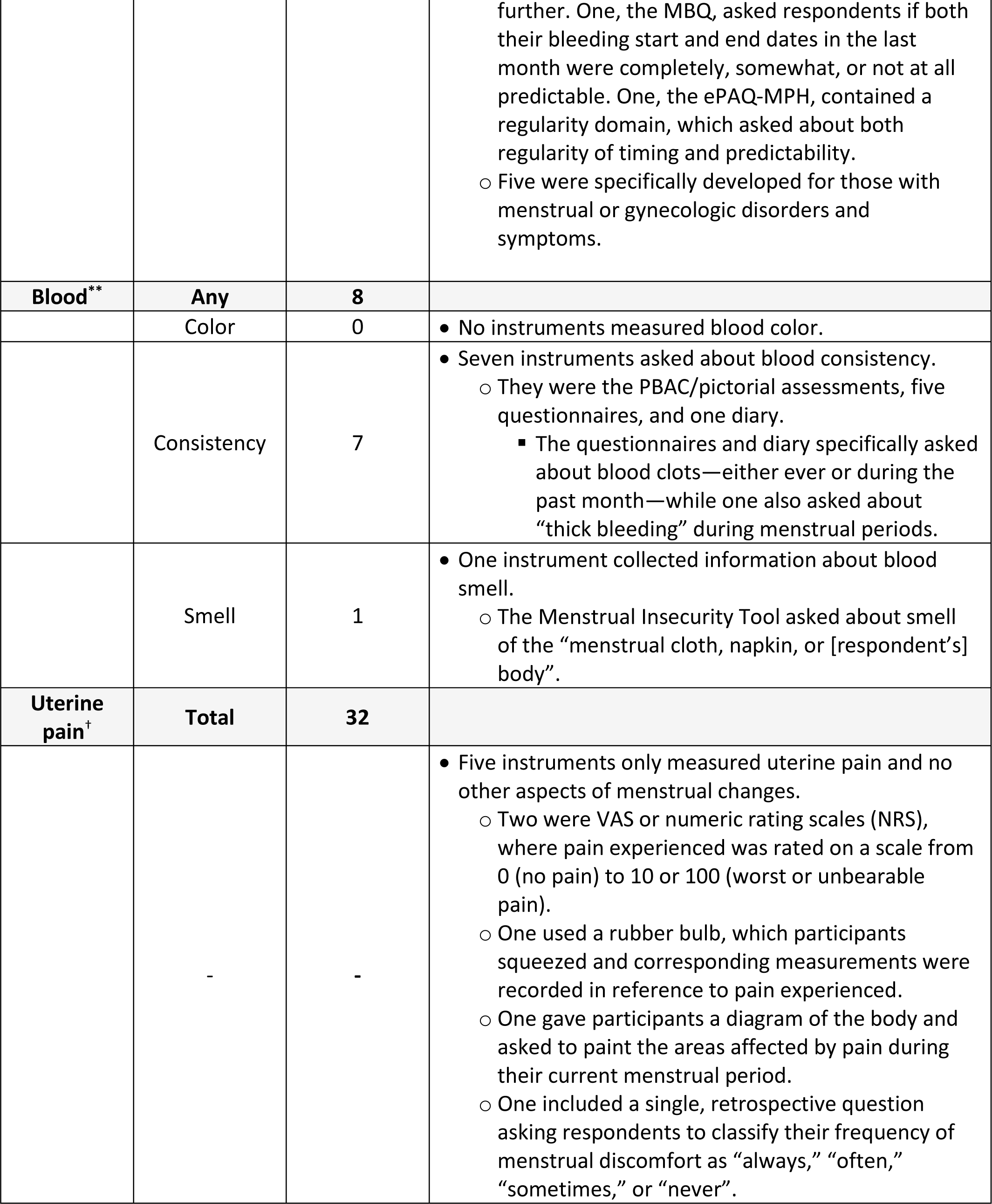
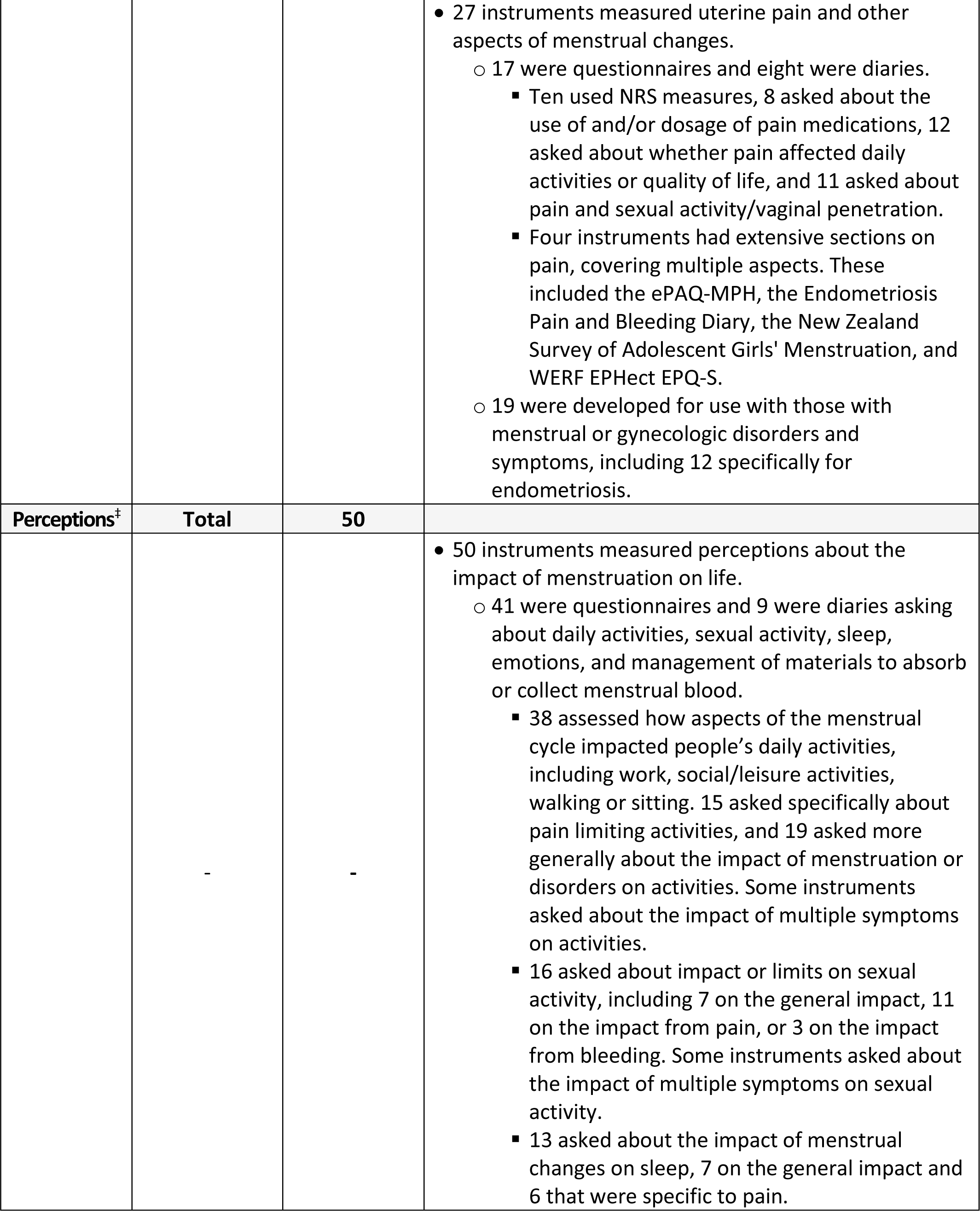
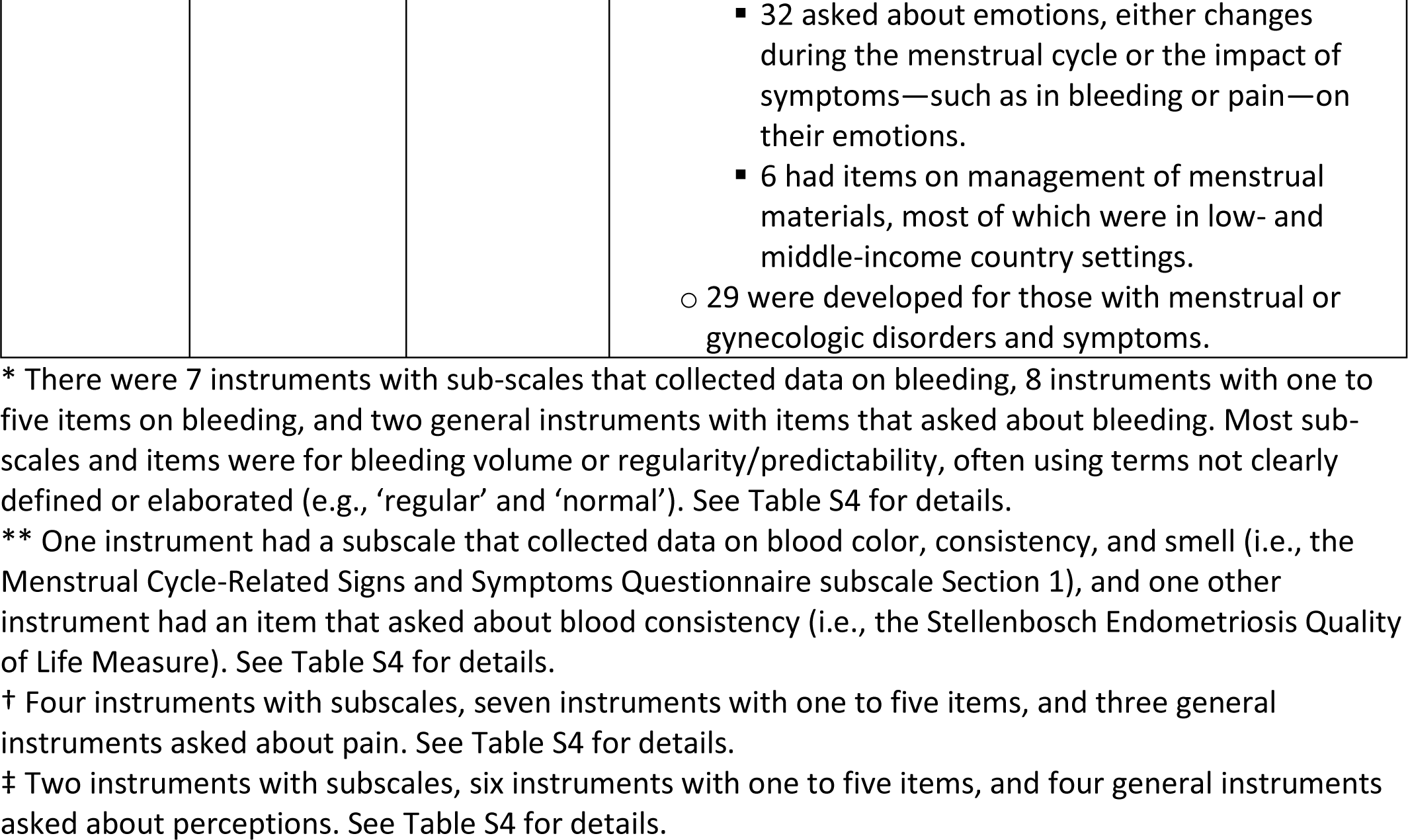
How full instruments measured aspects of menstrual changes, including bleeding, blood, uterine pain, and perceptions.

| Aspect of menstrual changes | Parameter | Number of instruments | How instruments measured |
| --- | --- | --- | --- |
| Bleeding* | Any | 33 |  |
|  | Duration | 12 | • One instrument only measured bleeding duration and no other parameter of bleeding or other aspects of menstrual changes.<br>○ It used prospective diaries to record the first and last days of menses/bleeding episodes just to measure duration.<br>• 11 instruments measured bleeding duration and other aspects of menstrual changes.<br>○ Three measured duration and another parameter of bleeding, either using diaries and/or annual interviews or a question on days of bleeding for every menstrual period over four months.<br>○ Eight measured bleeding duration along with other menstrual changes (i.e., blood, pain, or perceptions).<br>▪ Seven were questionnaires generally asking respondents to note how many days their menses/bleeding episodes last on average, either in general or in the last three months. Three specifically asked if respondents had bleeding for more than seven days per month.<br>▪ One diary asked respondents to note if they had bleeding on specific days.<br>○ Six were developed for people with menstrual or gynecologic disorders and symptoms (e.g., HMB, endometriosis, or fibroids). |
|  | Volume | 26 | • Five instruments only measured bleeding volume and no other parameter of bleeding or other aspects of menstrual changes.<br>○ Three semi-quantitatively measured blood volume via used menstrual products, including alkaline hematin assays and menstrual collection or record and recall measures.<br>○ One relied on respondents to estimate bleeding volume through the Mansfield-Voda-Jorgensen Menstrual Bleeding Scale.<br>○ One was a statistical model for estimating blood loss that was developed based on previously collected hematological values, daily diaries, and patient age among participants with HMB.<br>• 21 instruments measured bleeding volume and other aspects of menstrual changes.<br>○ 14 were questionnaires, five were diaries, one used pictorial references, and one was a visual analog scale (VAS) where volume was rated on a scale from 0 (no bleeding) to 100 (the heaviest possible bleeding ever experienced).<br>▪ Most asked about perceived volume of blood loss, usually by asking respondents to describe their bleeding in some range of light, medium, or heavy and/or reporting on the number of menstrual products (pads and/or tampons) they used on the heaviest day of their period.<br>· Terms like 'light', 'heavy' and/or 'spotting' were not always or consistently defined across instruments, and there was a wide range for the frame of reference for recall, with diaries asking every day, other instruments asking about the last month or last menses/bleeding episode, and others asking more generally about experiences people typically have during menses/bleeding episodes.<br>▪ Some instruments also asked how many days of heavy bleeding the respondent experienced during the last cycle and how many days required double protection with multiple products at the same time. A few asked whether respondents had bleeding heavy enough to stain clothing or required getting up<br>in the middle of the night to change menstrual products.<br>▪ PBAC and other similar pictorial assessments had respondents estimate the amount of bleeding via pictorials of used pads and/or tampons.<br>○ 18 were designed for use by people with menstrual or gynecologic disorders and symptoms. |
|  | Frequency | 8 | • Three instruments only measured bleeding frequency and no other parameter of bleeding or other aspects of menstrual changes.<br>○ They asked respondents a few retrospective questions (i.e., “How long is your menstrual cycle, on average? In other words, how many days are there from the first day of one menstrual period to the first day of the next period?”) or to recall the first date of their last menstrual period. Another used a retrospective questionnaire on usual, shortest, and longest menstrual cycle length in the past 12 months, and this was compared to a prospective diary for two menses/bleeding episodes.<br>• Five instruments measured bleeding frequency and other aspects of menstrual changes.<br>○ One was a diary.<br>○ Four were questionnaires asking respondents to state how many days there were, on average, between the start or first day of one menses/bleeding episode to the first day of the next menses/bleeding episode, or asking whether their menstrual cycle was between 21 and 45 days. |
|  | Regularity/predictability | 9 | • No instruments only measured bleeding regularity/predictability without other parameters of bleeding or other aspects of menstrual changes.<br>• Nine instruments measured bleeding regularity/predictability and other aspects of menstrual changes.<br>○ All were questionnaires, generally asking respondents to report if their bleeding was “regular” or “irregular” in general or over the past three months, but regularity was not defined<br>further. One, the MBQ, asked respondents if both their bleeding start and end dates in the last month were completely, somewhat, or not at all predictable. One, the ePAQ-MPH, contained a regularity domain, which asked about both regularity of timing and predictability.<br>○ Five were specifically developed for those with menstrual or gynecologic disorders and symptoms. |
| **Blood\*\*** | **Any** | **8** |  |
|  | Color | 0 | • No instruments measured blood color. |
|  | Consistency | 7 | • Seven instruments asked about blood consistency.<br>○ They were the PBAC/pictorial assessments, five questionnaires, and one diary.<br>▪ The questionnaires and diary specifically asked about blood clots—either ever or during the past month—while one also asked about “thick bleeding” during menstrual periods. |
|  | Smell | 1 | • One instrument collected information about blood smell.<br>○ The Menstrual Insecurity Tool asked about smell of the “menstrual cloth, napkin, or [respondent’s] body”. |
| **Uterine pain†** | **Total** | **32** |  |
|  | - | - | • Five instruments only measured uterine pain and no other aspects of menstrual changes.<br>○ Two were VAS or numeric rating scales (NRS), where pain experienced was rated on a scale from 0 (no pain) to 10 or 100 (worst or unbearable pain).<br>○ One used a rubber bulb, which participants squeezed and corresponding measurements were recorded in reference to pain experienced.<br>○ One gave participants a diagram of the body and asked to paint the areas affected by pain during their current menstrual period.<br>○ One included a single, retrospective question asking respondents to classify their frequency of menstrual discomfort as “always,” “often,” “sometimes,” or “never”. |
|  |  |  | • 27 instruments measured uterine pain and other aspects of menstrual changes.<br>○ 17 were questionnaires and eight were diaries.<br>▪ Ten used NRS measures, 8 asked about the use of and/or dosage of pain medications, 12 asked about whether pain affected daily activities or quality of life, and 11 asked about pain and sexual activity/vaginal penetration.<br>▪ Four instruments had extensive sections on pain, covering multiple aspects. These included the ePAQ-MPH, the Endometriosis Pain and Bleeding Diary, the New Zealand Survey of Adolescent Girls' Menstruation, and WERF EPHeCT EPQ-S.<br>○ 19 were developed for use with those with menstrual or gynecologic disorders and symptoms, including 12 specifically for endometriosis. |
| **Perceptions<sup>‡</sup>** | **Total** | **50** |  |
|  | - | - | • 50 instruments measured perceptions about the impact of menstruation on life.<br>○ 41 were questionnaires and 9 were diaries asking about daily activities, sexual activity, sleep, emotions, and management of materials to absorb or collect menstrual blood.<br>▪ 38 assessed how aspects of the menstrual cycle impacted people's daily activities, including work, social/leisure activities, walking or sitting. 15 asked specifically about pain limiting activities, and 19 asked more generally about the impact of menstruation or disorders on activities. Some instruments asked about the impact of multiple symptoms on activities.<br>▪ 16 asked about impact or limits on sexual activity, including 7 on the general impact, 11 on the impact from pain, or 3 on the impact from bleeding. Some instruments asked about the impact of multiple symptoms on sexual activity.<br>▪ 13 asked about the impact of menstrual changes on sleep, 7 on the general impact and 6 that were specific to pain. |
|  |  |  | ▪ 32 asked about emotions, either changes during the menstrual cycle or the impact of symptoms—such as in bleeding or pain—on their emotions.<br>▪ 6 had items on management of menstrual materials, most of which were in low- and middle-income country settings.<br>○ 29 were developed for those with menstrual or gynecologic disorders and symptoms. |

\* There were 7 instruments with sub-scales that collected data on bleeding, 8 instruments with one to five items on bleeding, and two general instruments with items that asked about bleeding. Most sub-scales and items were for bleeding volume or regularity/predictability, often using terms not clearly defined or elaborated (e.g., ‘regular’ and ‘normal’). See Table S4 for details.
\*\* One instrument had a subscale that collected data on blood color, consistency, and smell (i.e., the Menstrual Cycle-Related Signs and Symptoms Questionnaire subscale Section 1), and one other instrument had an item that asked about blood consistency (i.e., the Stellenbosch Endometriosis Quality of Life Measure). See Table S4 for details.
† Four instruments with subscales, seven instruments with one to five items, and three general instruments asked about pain. See Table S4 for details.
‡ Two instruments with subscales, six instruments with one to five items, and four general instruments asked about perceptions. See Table S4 for details.

### Measure quality of full instruments

When assessing measure quality, we found only five of the 68 full instruments (7%) had data on each of the six attributes of measure quality (i.e., conceptual or measurement model, reliability, content validity, construct validity, responsiveness, and sensitive nature of questions). These were the PBAC, EHP-30, Dysmenorrhea Daily Diary, MBQ, and a quantitative model for menstrual blood loss [62], each indicated by †† in Table 4. All but three instruments (96%, n=65) had evidence of a conceptual or measurement model and most also included evidence of content validity (81%, n=55), construct validity (84%, n=57) and reliability (66%, n=45); however, less than a third of instruments had evidence on responsiveness (31%, n=21), and less than a fifth (19%, n=13) had evidence on question sensitivity (Figure 2).

**Fig. 2.**
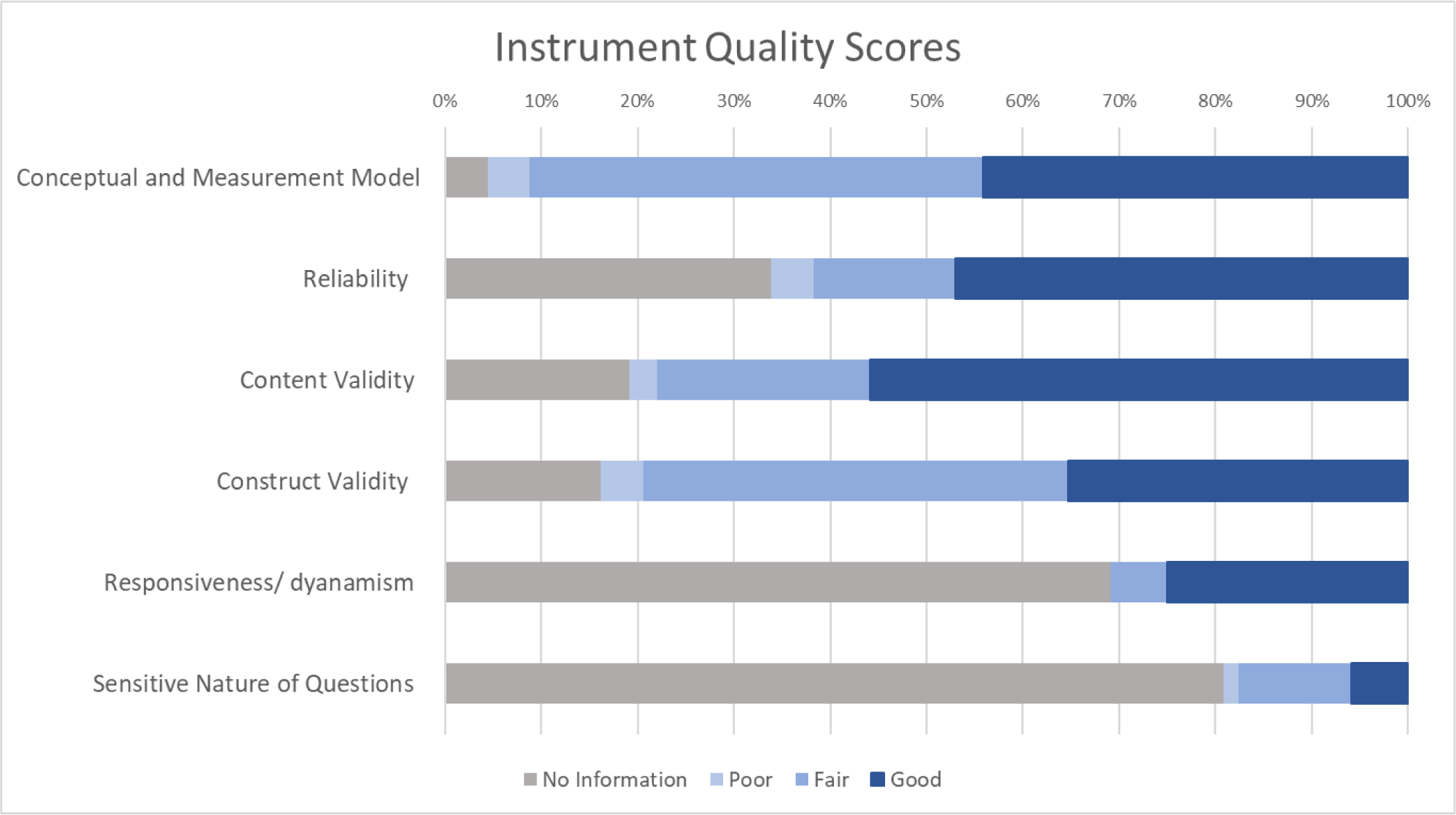
Instrument measure quality by attribute for full instruments.

Of the 68 full instruments, 18% (n=12) had an overall **good** measure quality score, about three quarters (74%, n=50) had a **fair** measure quality score, and 9% (n=6) had a **poor** measure quality score (Table 4 and Figure 2). When we looked at individual attributes of measure quality, over half of instruments had a good score for content validity (56%, n=38), 47% had a good score for reliability (n=32), 44% had a good score for conceptual or measurement model (n=30), and over a third of instruments (35%, n=24) had a good score for construct validity; however, only a quarter had a good score for responsiveness (25%, n=17), and only 4 instruments (6%) had a good score for question sensitivity.

### Utility for clinical trials of full instruments

When assessing clinical trial utility, we found 11 full instruments (16%) had data on each of the five attributes of utility (i.e., interpretability of results, transferability, participant burden, and investigator burden), each indicated by ‡ in Table 4. All but three instruments (96%) had information on participant burden, 84% (n=57) had evidence of the interpretability of the instrument results, and slightly less than two thirds (60%, n=41) had documented investigator burden; however, only just over one third (37%, n=25) had evidence of transferability (Figure 3).

**Fig. 3.**
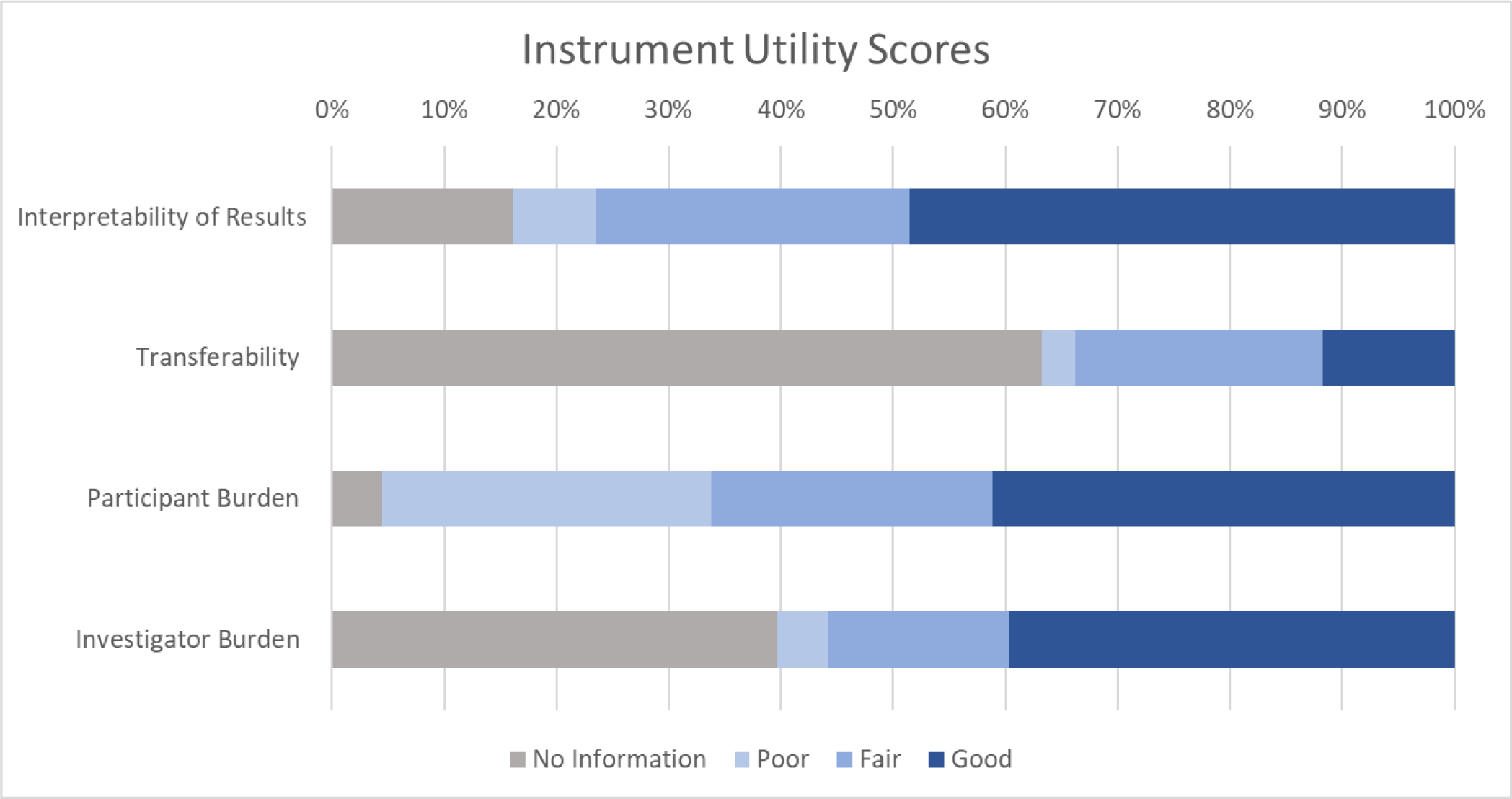
Instrument utility in clinical trials by attribute for full instruments.

Of the 68 full instruments, 22% (n=15) had an overall **good** clinical trial utility score, almost two thirds (62%, n=42) had a **fair** score, and 13% (n=9) had a **poor** score (Table 4 and Figure 3). When we looked at individual attributes of clinical trial utility, almost half of instruments (49%, n=33) had a good score for the interpretability of results, about 40% had good scores for participant burden (41%, n=28) or investigator burden (40%, n=27), but only 8 instruments (12%) had good scores for transferability.

### Overall full instrument evidence

Only the PBAC had evidence on all attributes of measure quality and all attributes of clinical trial utility, and only three instruments had both a good measure quality score and a good clinical trial utility score: EHP-5, the Spanish Society of Contraception Quality-of-Life (SEC-QOL), and the SAMANTA Questionnaire. Thirteen instruments had both measure quality scores and clinical trial utility scores greater than 2.5. Only one instrument, the Squeezing Pain Bulb, had both poor measure quality and poor clinical trial utility. Full instrument total evidence scores ranged from 4 for the World Health Organization Disability Assessment Schedule 2.0 to 332 for the EHP-30, with an overall median score across instruments of 16 and mean score of 27 (Table 4). Overall, the following instruments had the five highest scores across measure quality, clinical trial utility, and total evidence: EHP-30, EHP-5, UFS-QOL, PBAC, and MBQ.

## DISCUSSION

Our broad, transdisciplinary systematic review on the measurement of menstrual changes caused by any intrinsic or extrinsic factor, etiology, or source yielded 174 relevant articles and 94 instruments. Through our data extraction and analysis of these articles and instruments, we found several strengths and notable gaps in this literature around geographic and linguistic representation, how menstrual changes were measured, measure quality and clinical trial utility, and menstrual stigma, among others.

### Geographic and linguistic representation

We identified articles from all geographic regions and 50 countries, and full instruments in 28 languages, including over a quarter in more than one language. Despite this evidence of the breadth of the literature, three quarters of articles were from North America or Europe and almost half were from just the United States and United Kingdom. In addition, over half of full instruments were only in English, Spanish, French, or Portuguese. These findings indicate the existing instrument landscape centers around the US and Western Europe, as well as colonial languages.

### How menstrual changes were measured

We again found promising strengths mixed with important gaps when examining the menstrual changes instruments measured and how they were measured. Although many full instruments measured perceptions and at least one parameter of bleeding or pain, only 8 full instruments measured blood. It is possible this lack of data collection on blood is due to the wide influence of menstrual stigma, especially the common perspective that menstrual blood is ‘dirty’ and requires ‘hygiene’ products to cleanse, absorb, and hide blood or odor [52,63,64]. No full instruments measured all parameters for each of the four aspects of menstrual changes we assessed, and only 7 instruments measured at least one parameter for all four aspects. In addition, across all aspects of menstrual changes, we did not find high levels of uniformity between instruments regarding how they measured each menstrual change, and many did not explain or define key terms (e.g., ‘heavy’, ‘regular’), leaving their interpretation up to each respondent. This lack of clarity and specificity raises concerns about measurement error for a topic like menstruation and the wider menstrual cycle, around which there is high stigma and low health literacy and therefore, reduced shared understanding and references. These findings indicate there is a lack of instruments that examine all parameters and aspects of changes to the menstrual cycle in a comprehensive and standardized way.

Nearly 60% of full instruments we identified were developed for those with menstrual or gynecologic disorders and symptoms. In fact, the 3 instruments that accounted for almost a quarter of all identified articles—the EHP-30, PBAC, and USF-QOL—were each developed for use in populations with endometriosis, HMB, and fibroids, respectively. Instruments for these populations are of crucial importance, and it is encouraging to see over 70% of identified articles published in the last 5 years examine menstrual or gynecologic disorders and symptoms. However, the measurement of menstrual changes resulting from these disorders, such as very heavy bleeding and high levels of pain, may not translate to the menstrual changes experienced by the wider menstruating population or to the range of menstrual changes likely to occur across clinical trials and related research. For example, the extension of an instrument developed for those with HMB to a clinical trial of a hormonal contraceptive—which generally decreases bleeding volume—is yet to be supported by evidence. This difference is important because we could hypothesize, for example, there would be a difference in recall from a bleeding episode that resulted in stained clothing (i.e., from HMB) compared to a bleeding episode that did not interfere with daily activities (i.e., from a hormonal contraceptive). Because of these findings, instruments likely need to be developed or modified to capture a wider array of changes in bleeding, blood, and pain, as well as changes that are of smaller—but still meaningful—magnitude.

### Instrument quality and utility

From our assessments of measure quality and clinical trial utility for full instruments, we also found variability in our outcomes. Over 80% of instruments had either fair or good scores for measure quality or clinical trial utility, and only one had both poor measure quality and poor clinical trial utility. On the other hand, only three instruments had both good measure quality and good clinical trial utility.

We also note almost all instruments had evidence supporting some quality and utility attributes but not others. Sixty percent or more of instruments had evidence of a conceptual or measurement model, reliability, content validity, or construct validity for measure quality, or had evidence of interpretability of results, participant burden, or investigator burden for clinical trial utility; almost a quarter of instruments had evidence of each of these seven attributes. On the other hand, only one instrument— the PBAC—had evidence for all attributes of quality and utility, and over 60% of instruments did not have evidence of responsiveness, question sensitivity, or transferability, with nearly 40% not having evidence of any of the three. Each of these largely missing attributes are likely to be important for any instrument used broadly, especially in clinical trials. Such an instrument will need to: (a) capture changes during investigational drug use (responsiveness); (b) not be viewed as too intrusive or stigmatizing (question sensitivity); and (c) be used in multiple linguistic and sociocultural contexts (transferability).

### Menstrual stigma and other notable gaps

Our findings on the limited measurement for blood and lack of evidence for question sensitivity highlight the importance of menstrual stigma. We often found a contradiction during the development and validation of instruments; although menstrual stigma was frequently acknowledged as part of the sociocultural milieu surrounding menstruation, instruments generally did not adequately address menstrual stigma or how stigma may relate to question sensitivity and the potential impact of this on data quality or measurement error.

Beyond the difficulty of measurement due to menstrual stigma, there is innate complexity in measuring changes to a biological process that, itself, consists of so many facets that change over time and vary between individuals [65,66]. For example, there are changes between days of a single menstrual cycle (e.g., different bleeding and/or pain experienced on different days of a cycle), differences among menstrual cycles during the same year, and shifts over the menstruating life course for one individual person who menstruates, as well as a multitude of differences between people [67–69]. These factors are important when we consider just under half of articles for the identified full instruments had cross-sectional study designs. In fact, this study design limitation could be the reason we found a lack of evidence on instrument responsiveness and measurement of more temporally-related parameters like bleeding frequency and regularity/predictability.

In addition to the gaps in the literature and instrument landscape already mentioned, three additional findings warrant attention. First, only just over a third of instruments used electronic data collection. Although this may be partly due to our review extending through 2006, new and refined instruments should strongly consider this approach given the data quality and monitoring benefits of electronic data collection and with the current proliferation of period tracking and other FemTech applications [70,71]. In addition, there is a need to establish the equivalence between existing paper instruments and any electronic versions developed, ideally in accordance with established approaches like the International Professional Society for Health Economics and Outcomes Research (ISPOR) good research practices on use of mixed mode PROs [72].

Second, there is a lack of attention paid to the two ends of the menstruating life course. There were only ten instruments specifically developed with data from adolescents and three instruments developed for those in perimenopause, both groups who can experience an increased amount of variability and change in their menstrual cycles as compared to the middle of the menstruating years [73]. In addition, data on older menstruators were often collapsed for people who were in perimenopause and menopause/post-menopause, or age was commonly used as a proxy for this process and transition. Although the age range for menopause is narrower than that of menarche, given the general lack of research around menopause and the preceding and succeeding years, it seems the opposite should be true (i.e., more data and larger sample sizes among people around the end of their menstruating years is warranted) [74].

Third, we found a lack of inclusion for trans and gender nonbinary populations in all articles for all instruments. As we note in the introduction of this paper, people who menstruate may or may not identify as women or girls, and not all women and girls menstruate. It is important to engage all populations who menstruate in the development of instruments to measure changes to the menstrual cycle. Inclusion of sexual and gender minority (SGM) individuals who menstruate in clinical trials is a noted priority among NIH and other funders and researchers. In addition to NIH establishing its SGM Research Office in 2015, *clinical research* is the first theme of the current Strategic Plan to Advance Research on the Health and Well-being of SGM populations [75].

### Limitations of the review

Although we followed PRISMA guidelines and included ‘inter-reviewer reliability’ checks, weekly meetings, and multiple reviewers per article, there are a few limitations to note about our review process. The most important limitations are related to decisions made regarding the scope of the review to make it focused and feasible. First, we only included four aspects of changes to the menstrual cycle: changes in bleeding, blood, pain, and perceptions of bleeding, blood, or pain. Although these aspects are likely the most studied thus far, there are many other important changes to the menstrual cycle, including in hormone levels, the phases of the menstrual cycle, characteristics of those phases, and other symptoms besides pain. As the study of menstrual health grows, it will be important for future reviews to consider these areas of research. We also limited our scope to the menstrual cycle, excluding other types of uterine bleeding, such as bleeding during pregnancy, while breastfeeding, and after menopause. Future insights into how these types of bleeding relate to bleeding during the menstrual cycle will be important to the research and understanding of all uterine bleeding.

We also note a few limitations related to our review process. First, although all authors have training and experience across multiple disciplines, none are experts in all fields from which we drew our literature given our transdisciplinary approach. We aimed to address this limitation by consulting other experts internally at FHI 360 and members of a related global task force when we encountered a question or issue outside of our knowledgebase, but it is still possible we missed articles, data for extraction, or other elements due to this limitation. Second, the primary impetus for the review among the authors was to inform measurement of menstrual changes in the context of contraceptive clinical trials, so we cannot completely rule out the possibility this internal aim may have influenced our decisions about including or excluding articles. From the very beginning of the review, however, we sought for the review to be useful across contexts and disciplines, so our protocol and process were designed and implemented with that purpose in mind. Third, we may have missed articles by deciding to not include the CINAHL and PsycINFO databases in addition to those we did include (i.e., MEDLINE, Embase, and the instrument databases). Despite reviewing at least 50 articles most relevant to search strategies for CINAHL and PsycINFO and finding none aligned with our inclusion criteria, it is possible there were articles relevant to our review in the rest of the search results from these two databases. Fourth, because we did not want to exclude articles from any region or language but are not fluent in all languages, we used Google Translate for some screening and review. It is, therefore, possible this translation did not allow us to sufficiently evaluate articles per our inclusion and exclusion criteria. For the two relevant articles not written in English, we did complete data extraction with a fluent colleague. Overall, there may be additional limitations about which we are not aware that may have biased the results of our systematic review. Our hope is, however, we took steps to mitigate as many as possible by having our protocol and instrument evaluation criteria reviewed by other experts, following best practice guidelines, and taking steps to reduce individual variability and biases.

## CONCLUSION

Despite the novel, broad, and transdisciplinary approach to our systematic review, the current instrument landscape, limitations in the literature, and gaps in evidence on measure quality and clinical trial utility indicate there is a need to examine changes to the menstrual cycle in a more complete, inclusive, and standardized way. Rigorous formative research—across sociocultural contexts—that is focused on how all people who menstruate experience and understand their menstrual cycles and more fully addresses menstrual stigma can inform the development of new or modified instruments to meet this need. We also identified a need for greater evidence of the validity for existing and new instruments. For the clinical trial context, FDA guidance on selecting, developing, or modifying patient-reported outcomes like menstrual changes indicate there must be evidence to support the use of an instrument for the specific concepts of interest and context of use [37]. At a minimum, per this guidance, evidence would be needed to support the use of the instruments identified and assessed in this review in the clinical trial context with a broader patient population (i.e., context of use) and to measure the full scope of menstrual changes that people experience (i.e., concept of interest). In addition, the recent emergence of core outcome sets within areas like HMB and endometriosis will be useful to promote standardization of validated instruments, especially if these efforts are interdisciplinary and coordinated across research areas.

The findings of our review will be helpful in developing new or modified instruments that assess menstrual changes in a validated, comprehensive way. If used across the many fields that study menstrual health, data from these standardized instruments can contribute to an interdisciplinary, systemic, and holistic understanding of menstruation and the menstrual cycle. In turn, this improved understanding can be translated into ways to enhance the health and wellbeing of people who menstruate.

## Data Availability

All data produced in the present work are contained in the manuscript.

## ACKNOWLEDGEMENTS

The authors would like to thank Laneta Dorflinger, Rebecca Callahan, and members of the Global Contraceptive Induced Menstrual Changes (CIMC) Task Force for their feedback on the draft review protocol. We are very grateful for the guidance and assistance provided by the FHI 360 health sciences library throughout the development and implementation of our literature search strategy (Allison Burns and Carol Manion) and during retrieval of full text articles (Tamara Fasnacht). We would also like to acknowledge Betsy Costenbader, Kavita Nanda, and Global CIMC Task Force members for their feedback on the draft data extraction forms, and Kavita Nanda for clinical advice. We would also like to thank Valeria Bahamondes for her assistance in the full text review and data extraction of the two Portuguese papers that were included. The authors also appreciate Laneta Dorflinger and Kate McQueen for their review of and feedback on drafts of this manuscript.

## SUPPORTING INFORMATION

**S1 Table.** Preferred Reporting Items for Systematic Reviews and Meta-Analysis (PRISMA) checklist

**S2 Appendix**. Details on Review Methods.

**S3 Table.** All articles included after title/abstract screening and full text review.

**S4 Table.** Characteristics of sub-scales, items, and general instruments.

