## Supporting Information (S1-S4) for "Measurement of changes to the menstrual cycle: A transdisciplinary systematic review evaluating measure quality and utility for clinical trials"

#### S1 Table. Preferred Reporting Items for Systematic Reviews and Meta-Analysis (PRISMA) checklist.

##### PRISMA 2020 Checklist

| Section and Topic | Item # | Checklist item | Location where item is reported |
|---|---|---|---|
| **TITLE** | | | |
| Title | 1 | Identify the report as a systematic review. | Pg 1 |
| **ABSTRACT** | | | |
| Abstract | 2 | See the PRISMA 2020 for Abstracts checklist. | Pg 2 |
| **INTRODUCTION** | | | |
| Rationale | 3 | Describe the rationale for the review in the context of existing knowledge. | Pg 5-8 |
| Objectives | 4 | Provide an explicit statement of the objective(s) or question(s) the review addresses. | Pg 8 |
| **METHODS** | | | |
| Eligibility criteria | 5 | Specify the inclusion and exclusion criteria for the review and how studies were grouped for the syntheses. | Pg 9-10, Supplementary Appendix S2 |
| Information sources | 6 | Specify all databases, registers, websites, organisations, reference lists and other sources searched or consulted to identify studies. Specify the date when each source was last searched or consulted. | Pg 8-9, Supplementary Appendix S2 |
| Search strategy | 7 | Present the full search strategies for all databases, registers and websites, including any filters and limits used. | Table 1, Supplementary Appendix S2 |
| Selection process | 8 | Specify the methods used to decide whether a study met the inclusion criteria of the review, including how many reviewers screened each record and each report retrieved, whether they worked independently, and if applicable, details of automation tools used in the process. | Pg 10-11, Supplementary Appendix S2 |
| Data collection process | 9 | Specify the methods used to collect data from reports, including how many reviewers collected data from each report, whether they worked independently, any processes for obtaining or confirming data from study investigators, and if applicable, details of automation tools used in the process. | Pg 10-15 ,Supplementary Appendix S2 |
| Data items | 10a | List and define all outcomes for which data were sought. Specify whether all results that were compatible with each outcome domain in each study were sought (e.g. for all measures, time points, analyses), and if not, the methods used to decide which results to collect. | Pg 11-15, Supplementary Appendix S2 |
| | 10b | List and define all other variables for which data were sought (e.g. participant and intervention characteristics, funding sources). Describe any assumptions made about any missing or unclear information. | Pg 11-15, Supplementary Appendix S2 |
| Study risk of | 11 | Specify the methods used to assess risk of bias in the included studies, including details of the tool(s) | Pg 11-15, |

| Section and Topic | Item # | Checklist item | Location where item is reported |
|---|---|---|---|
| bias assessment | | used, how many reviewers assessed each study and whether they worked independently, and if applicable, details of automation tools used in the process. | Supplementary Appendix S2 |
| Effect measures | 12 | Specify for each outcome the effect measure(s) (e.g. risk ratio, mean difference) used in the synthesis or presentation of results. | NA |
| Synthesis methods | 13a | Describe the processes used to decide which studies were eligible for each synthesis (e.g. tabulating the study intervention characteristics and comparing against the planned groups for each synthesis (item #5)). | Pg 11-15, Supplementary Appendix S2 |
| | 13b | Describe any methods required to prepare the data for presentation or synthesis, such as handling of missing summary statistics, or data conversions. | Pg 11-15, Supplementary Appendix S2 |
| | 13c | Describe any methods used to tabulate or visually display results of individual studies and syntheses. | Pg 11-15, Supplementary Appendix S2 |
| | 13d | Describe any methods used to synthesize results and provide a rationale for the choice(s). If meta-analysis was performed, describe the model(s), method(s) to identify the presence and extent of statistical heterogeneity, and software package(s) used. | Pg 11-15, Supplementary Appendix S2 |
| | 13e | Describe any methods used to explore possible causes of heterogeneity among study results (e.g. subgroup analysis, meta-regression). | NA |
| | 13f | Describe any sensitivity analyses conducted to assess robustness of the synthesized results. | NA |
| Reporting bias assessment | 14 | Describe any methods used to assess risk of bias due to missing results in a synthesis (arising from reporting biases). | Pg 11-15, Supplementary Appendix S2 |
| Certainty assessment | 15 | Describe any methods used to assess certainty (or confidence) in the body of evidence for an outcome. | Pg 11-15, Supplementary Appendix S2 |
| **RESULTS** | | | |
| Study selection | 16a | Describe the results of the search and selection process, from the number of records identified in the search to the number of studies included in the review, ideally using a flow diagram. | Pg 16-17, Figure 1 |
| | 16b | Cite studies that might appear to meet the inclusion criteria, but which were excluded, and explain why they were excluded. | Pg 16-17 |
| Study characteristics | 17 | Cite each included study and present its characteristics. | Supplementary Table S3 |
| Risk of bias in studies | 18 | Present assessments of risk of bias for each included study. | Pg 25-27, Figure 2-3 |

| Section and Topic | Item # | Checklist item | Location where item is reported |
|---|---|---|---|
| Results of individual studies | 19 | For all outcomes, present, for each study: (a) summary statistics for each group (where appropriate) and (b) an effect estimate and its precision (e.g. confidence/credible interval), ideally using structured tables or plots. | NA |
| Results of syntheses | 20a | For each synthesis, briefly summarise the characteristics and risk of bias among contributing studies. | Pg 17-27 |
| | 20b | Present results of all statistical syntheses conducted. If meta-analysis was done, present for each the summary estimate and its precision (e.g. confidence/credible interval) and measures of statistical heterogeneity. If comparing groups, describe the direction of the effect. | NA |
| | 20c | Present results of all investigations of possible causes of heterogeneity among study results. | NA |
| | 20d | Present results of all sensitivity analyses conducted to assess the robustness of the synthesized results. | NA |
| Reporting biases | 21 | Present assessments of risk of bias due to missing results (arising from reporting biases) for each synthesis assessed. | Pg 25-27, Figure 2-3 |
| Certainty of evidence | 22 | Present assessments of certainty (or confidence) in the body of evidence for each outcome assessed. | Pg 25-27, Figure 2-3 |
| **DISCUSSION** | | | |
| Discussion | 23a | Provide a general interpretation of the results in the context of other evidence. | Pg 27-31 |
| | 23b | Discuss any limitations of the evidence included in the review. | Pg 31-33 |
| | 23c | Discuss any limitations of the review processes used. | Pg 31-33 |
| | 23d | Discuss implications of the results for practice, policy, and future research. | Pg 33-34 |
| **OTHER INFORMATION** | | | |
| Registration and protocol | 24a | Provide registration information for the review, including register name and registration number, or state that the review was not registered. | Pg 8 |
| | 24b | Indicate where the review protocol can be accessed, or state that a protocol was not prepared. | Pg 8 |
| | 24c | Describe and explain any amendments to information provided at registration or in the protocol. | NA |
| Support | 25 | Describe sources of financial or non-financial support for the review, and the role of the funders or sponsors in the review. | With PLOS submission |
| Competing interests | 26 | Declare any competing interests of review authors. | With PLOS submission |
| Availability of data, code and other materials | 27 | Report which of the following are publicly available and where they can be found: template data collection forms; data extracted from included studies; data used for all analyses; analytic code; any other materials used in the review. | Supplementary Appendix S2 |

*From:* Page MJ, McKenzie JE, Bossuyt PM, Boutron I, Hoffmann TC, Mulrow CD, et al. The PRISMA 2020 statement: an updated guideline for reporting systematic reviews. BMJ 2021;372:n71. doi: 10.1136/bmj.n71 For more information, visit: http://www.prisma-statement.org/

### Supplementary Appendix S2: Details on Review Methods

#### Systematic review protocol

We developed a protocol per PRISMA guidance, and protocol drafts were reviewed by experts in the fields of menstruation and contraception who are members of the Global Contraceptive-Induced Menstrual Changes (CIMC) Task Force [1]. We registered our review protocol in PROSPERO (ID: CRD42023420358) [2].

#### Search strategy

We conducted a multi-stage literature search in collaboration with the FHI 360 health sciences library to identify peer reviewed articles examining instruments to measure menstrual changes. First, we conducted preliminary searches in MEDLINE to refine our search strategy, including PubMed search terms recommended by the Consensus-based Standards for the selection of health Measurement Instruments (COSMIN) [3]. We then reviewed the 50 most relevant hits from the Embase, CINAHL, and PsycINFO databases to determine which should be included in our search strategy in addition to MEDLINE. Only Embase contained relevant articles within those 50 most relevant hits, so it was the only other database included in our final search. Table A shows the final search strategy for MEDLINE, which included largely Medical Subject Headings (MeSH) Major Topic terms and title or abstract search terms. The MEDLINE search strategy was adapted by an FHI 360 health sciences librarian for Embase (Table A). Final searches of MEDLINE and Embase were conducted, and the resulting records were uploaded into Covidence [4].

##### Table A. Search strategies

| Database | Search strategy | Date searched |
|---|---|---|
| MEDLINE | ("menstrual cycle"[MeSH Major Topic] OR "menstruation disturbances"[MeSH Major Topic] OR "Endometriosis"[MeSH Major Topic] OR "Uterine Diseases"[MeSH Major Topic] OR "menstrua*"[Title/Abstract] OR "menses"[Title/Abstract] OR "uterine bleeding"[Title/Abstract] OR "vaginal bleeding"[Title/Abstract] OR "amenorrhea"[Title/Abstract] OR "dysmenorrhea"[Title/Abstract] OR "menorrhagia"[Title/Abstract] OR "oligomenorrhea"[Title/Abstract] OR "metrorrhagia"[Title/Abstract] OR | Original:<br>June 23, 2022<br><br>Updated:<br>October 5, 2023 |

| | | |
|---|---|---|
| | "hypermenorrhea"[Title/Abstract] OR "hypomenorrhea"[Title/Abstract] OR "polymenorrhea"[Title/Abstract])<br>AND<br>("Surveys and Questionnaires"[MeSH Major Topic] OR "Pain Measurement"[MeSH Major Topic] OR "Patient Reported Outcome Measures"[MeSH Major Topic] OR "psychometrics"[MeSH Major Topic] OR "Sensitivity and Specificity"[MeSH Major Topic] OR "Validation Study"[Publication Type] OR "Validation Studies as Topic"[MeSH Major Topic] OR "measur*"[Title] OR "method*"[Title] OR "questionnaire*"[Title] OR "scale"[Title] OR "tool*"[Title] OR "patient reported outcome measure*"[Title/Abstract] OR "psychometr*"[Title/Abstract])<br>AND<br>("2006/01/01"[Date - Publication] : "2023/10/05"[Date - Publication]) | |
| Embase | ('menstrual cycle'/exp/mj OR 'menstruation disorder'/exp/mj OR 'endometriosis'/exp/mj OR 'uterus disease'/exp/mj OR 'menstrua*':ab,ti OR 'menses':ab,ti OR 'uterine bleeding':ab,ti OR 'vaginal bleeding':ab,ti OR 'amenorrhea':ab,ti OR 'dysmenorrhea':ab,ti OR 'menorrhagia':ab,ti OR 'oligomenorrhea':ab,ti OR 'metrorrhagia':ab,ti OR 'hypermenorrhea':ab,ti OR 'hypomenorrhea':ab,ti OR 'polymenorrhea':ab,ti) AND ('measurement'/exp/mj OR 'questionnaire'/exp/mj OR 'pain measurement'/exp/mj OR 'patient-reported outcome'/exp/mj OR 'psychometry'/exp/mj OR 'sensitivity and specificity'/exp/mj OR 'validation study'/exp/mj OR 'measur*':ti OR 'method*':ti OR 'questionnaire*':ti OR 'scale':ti OR 'tool':ti OR 'patient reported outcome measure*':ti,ab OR 'psychometr*':ti,ab) AND [2006-2023]/py AND [embase]/lim NOT [medline]/lim | Original:<br>June 28, 2022<br><br>Updated:<br>October 5, 2023 |
| NIH Common Data Element (CDE) Repository | Searched menstruation-related pre-defined topic areas:<br>• “menstruation scale”<br>• “menstrual period regularity type”<br>• “irregularity of menstrual cycle”<br>• “menstrual cycle typical days PhenX”<br>• “menstrual period last date”<br>• “menstrual period occurrence indicator” (Oct 2023 search) | Original:<br>October 11, 2022<br><br>Updated:<br>October 11, 2023 |

| | | |
|---|---|---|
| COSMIN[i] | Keyword search for relevant instruments containing "menstru*" or "bleed*" in title | Original<br>October 11, 2022<br><br>Updated<br>October 11, 2023 |
| COMET[ii] | Searched pre-defined Disease Names categories:<br>• "Abnormal uterine bleeding"<br>• "Endometriosis"<br>• "Endometriosis-related pain"<br>• "Heavy menstrual bleeding"<br>• "Uterine fibroids" | Original<br>October 11, 2022<br><br>Updated<br>October 11, 2023 |
| ePROVIDE | Keyword search for relevant instruments tagged "menstru*", "dysmenorrhea", or "menorrhagia" | Original<br>October 11, 2022<br><br>Updated<br>October 11, 2023 |

Next, we searched four instrument databases for any relevant instruments measuring menstrual changes: (a) the NIH Common Data Element (CDE) Repository [5], (b) the COSMIN database of systematic reviews of outcome measurement instruments [6], (c) the Core Outcome Measures in Effectiveness Trials (COMET) Database [7], and (c) ePROVIDE databases [8]. We detail search strategies for these instrument databases in Table A. Articles for any relevant instruments identified via these databases were uploaded into Covidence. We also planned to include instruments identified from searches of ClinicalTrials.gov and the Patient-Reported Outcomes Measurement Information System (PROMIS) database of measures, but multiple search strategies did not yield results we could screen and include.

Following screening and review of articles from the two literature databases (i.e., MEDLINE and Embase) and the four instrument databases (i.e., NIH CDE, COSMIN, COMET, and ePROVIDE), we completed two additional steps: (a) we extracted primary articles published since 1980 from all relevant review articles identified from the literature and instrument databases; and (b) we identified any original development articles for instruments developed before 2006. These primary articles and original development articles

[i] Consensus-based Standards for the selection of health Measurement Instruments (COSMIN) database of systematic reviews of outcome measurement instruments
[ii] Core Outcome Measures in Effectiveness Trials

were then uploaded into Covidence for screening. Book chapters were excluded at this stage of screening.

Overall, our goal was to include all articles published on the (a) development, (b) validation, or (c) review of instruments since January 1, 2006. For instrument development or validation (a and b), we selected 2006 because the last major revision of standardized CIMC measurement in contraceptive clinical trials was published in 2007; therefore, that revision would encompass instruments developed or validated prior to 2006. For instruments reviewed (c), we selected 1980 as our date limit for extracting primary papers from identified reviews because the initial efforts to standardize CIMC measurement in contraceptive clinical trials, led by the World Health Organization (WHO), were in the 1980s; therefore, that WHO work would already encompass literature before 1980.

#### Updated Search

After completing our systematic review, we conducted an updated search in October 2023 to ensure the results reported up-to-date findings. Original literature database searches (i.e., PubMed and Embase) covered January 1, 2006 through June 23, 2022, and updated searches covered June 23, 2022 through October 5, 2023. Original database searches (i.e., NIH CDE, COSMIN, COMET, and ePROVIDE) were conducted on October 11, 2022 and updates on October 11, 2023. For all identified articles in both searches, we completed the same search, screening, and review processes described in the main paper. The main paper reports on total results from all searches combined. Details on each search follow.

##### Original search

Our original database searches yielded a total of 7,189 articles, of which 7,135 were from literature databases and 54 from instrument databases. Covidence removed 154 duplicates and we excluded 6,761 articles during title/abstract screening. During full text review, we excluded 93 articles for study design, article type, or population, 26 for not measuring menstrual changes, and 9 for no validation. We also identified one additional duplicate and found 23 relevant review articles. From these review articles we extracted 640 primary articles, of which 35 remained after title/abstract screening and full text review. During data extraction, we identified 6 instruments for which we did not have the original development papers, because either they were developed before 2006 (i.e., our search strategy date limit; n=5) or had not been captured via our search strategy (n=1). Across all sources, our searches yielded 7,835 articles. We removed 315 duplicates, excluded 7,171 articles during title and abstract screening, and excluded 190 articles during full text review. In total, we identified 159 relevant full text articles of instruments developed, validated, or reviewed between January 1, 2006 and June 23, 2022. We present the PRISMA diagram for the original search in Figure A.

#### Updated search

Our original database searches yielded a total of 655 articles, of which 639 were from literature databases and 16 from instrument databases. Covidence removed 61 duplicates and we excluded 533 articles during title/abstract screening. During full text review, we excluded 22 articles for study design, article type, or population, 15 for not measuring menstrual changes, and 9 for no validation. We identified no relevant review articles, and no instruments for which we did not have the original development papers. In total, we identified 15 additional relevant full text articles of instruments developed, validated, or reviewed between June 23, 2022 and October 5, 2023. We present the PRISMA diagram for the updated search in Figure B.

The updated search yielded 15 additional articles on 11 full instruments (including 2 articles on one instruments, the EHP-30) and 3 broader instruments that included sub-scales (n=1) or a small number of items (n=2, both of which were not identified in the original search). Of the 11 new full instruments, 4 had not been identified in the original search (i.e., Pain Drawing, the World Health Organization Disability Assessment Schedule 2.0, the Bleeding and Pelvic Discomfort Scale, and the PERIOD-QOL).

#### Figure A: Original search PRISMA diagram

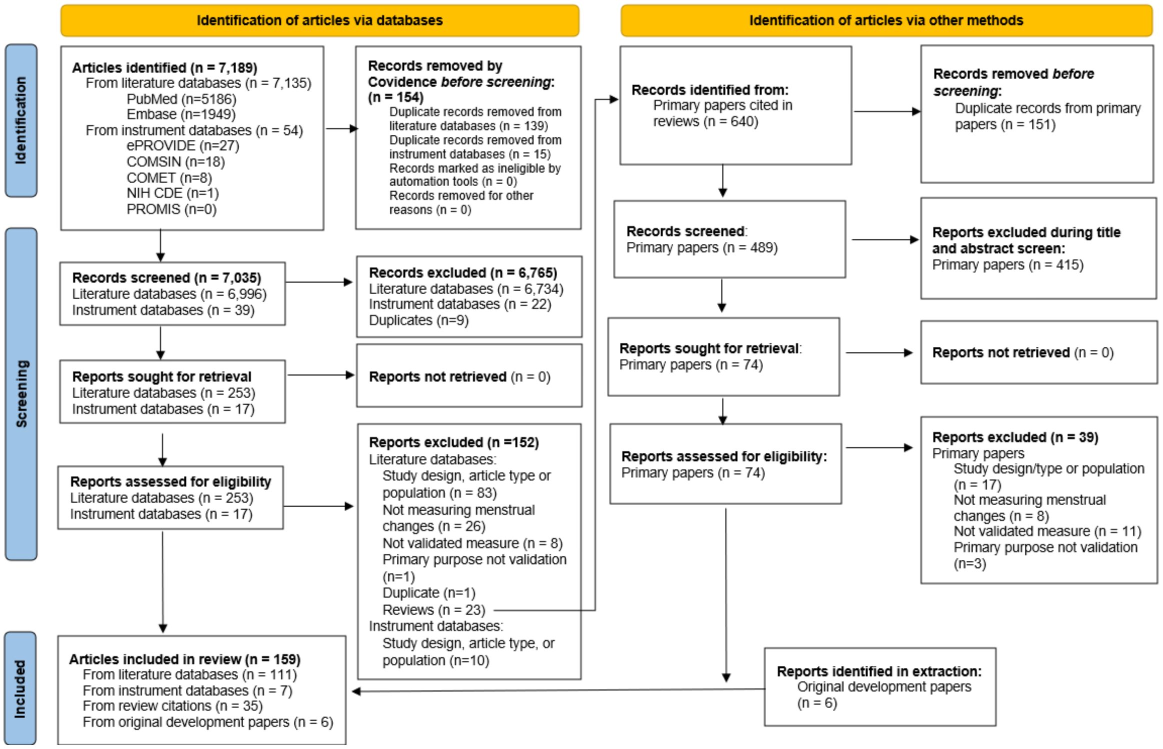


#### Figure B: Updated search PRISMA diagram

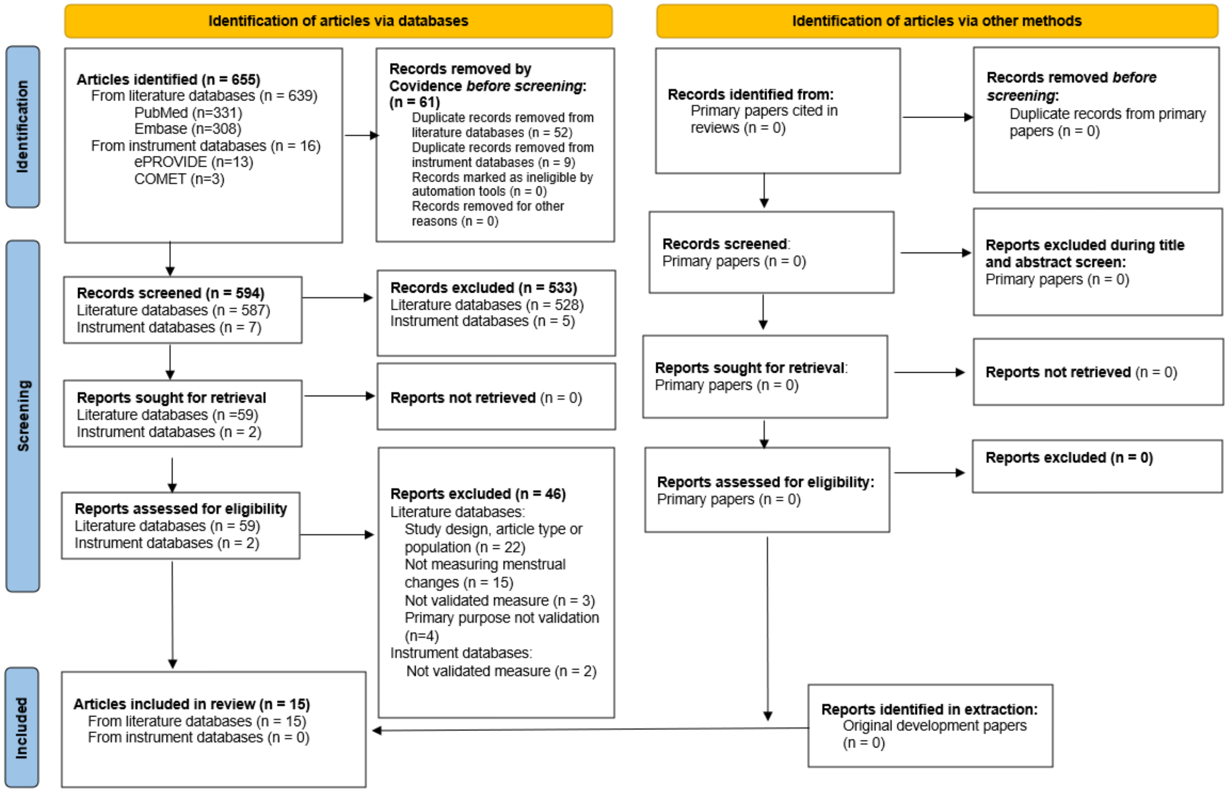

#### Inclusion/exclusion criteria

We included all peer-reviewed articles—including those with prospective, retrospective, or cross-sectional study designs, and review papers—that met our inclusion and did not meet our exclusion criteria. We detail these criteria in Table B, but briefly, we included articles that: (a) reported on the development or validation of instruments to measure menstrual changes, (b) used mixed methods or quantitative approaches, and (c) were published between January 1, 2006 and October 5, 2023. We did not impose any restrictions on article language, country, or geographic region. Articles using only qualitative methods and conference abstracts, editorials, and commentaries were excluded because they would not contain the information necessary to evaluate instrument quality and utility for clinical trials, per our second review question.

##### Table B: Inclusion and exclusion criteria

| | |
|---|---|
| Inclusion criteria | 1. Articles primarily focused on developing, validating, and/or evaluating instruments measuring menstrual changes or perceptions of menstrual changes, with information reported to assess instrument and/or study quality<br>2. Articles published between January 1, 2006 and October 5, 2023<br>3. Articles published in any language<br>4. Articles from any geographic region |
| Exclusion criteria | 1. Articles with only qualitative data<br>2. Articles that were conference abstracts, editorials, and commentaries<br>3. Articles whose primary purpose was noy validating instruments measuring menstrual change, such as studies focusing on biomarkers or biological pathways of menstrual changes, cancer screening instruments, or studies of social-behavioral correlates of menstrual changes<br>4. Articles reporting only on data from people in menopause |

Our definition of menstrual changes was adapted and broadened from the Global CIMC Task Force definition of changes to the menstrual cycle caused by contraception [1]. For the purposes of this review, the term, **menstrual changes**, includes four aspects (a) bleeding duration, volume, frequency, and/or regularity/predictability; (b) blood consistency, color, and/or smell; (c) pain or cramping; and (d) perceptions of bleeding, blood, or pain. We define perceptions as the perspectives on, attitudes about, experiences with, and acceptability of menstrual changes at the individual-level, interpersonal-level, community-level, and wider levels, including social norms. Examples of these four aspects of menstrual changes are: (a) an increase in how long bleeding lasts (bleeding duration), (b) a reduction of clotting (blood consistency), (c) a decrease in dysmenorrhea (pain), and (d) an impact on quality of life or attitudes (perceptions of changes).

We use the single term '**instrument**' to capture any measure, method, or approach to assess menstrual changes, including healthcare provider-reported, menstruator-reported, researcher-based, biomarker-based, or assay-based methods, and including those that may be deemed "objective" or "subjective" and both directly observable and personal perceptions of menstrual changes (adapted from [9]). Our definition of **development** or **validation** of instruments was intentionally broad, including any manner of validation or evaluation (e.g., reporting any evidence on validity, reliability, responsiveness, interpretability, and other attributes of measure quality or utility) and any development or validation informed by input from research participants who menstruate.

#### Developing data extraction forms and instrument evaluation

One author (SC) drafted the initial template data extraction form in Excel after input from the rest of the authors, and all authors reviewed and gave feedback on the draft data extraction form. The final data extraction form collected information in five areas: article information, study design and sample information, details on the instrument, measure quality attributes, and clinical trial utility attributes. Table C has details on the fields of the data extraction form for each of the five areas.

Table C: Fields of data extraction form.

| Information area | Information fields |
|---|---|
| 1. Article information | • Author initials<br>• Date extraction completed<br>• Covidence ID number<br>• First author<br>• Publication year<br>• Title |
| 2. Study design and sample information | • Region<br>• Country<br>• Language<br>• Sample size (analysis sample)<br>• Sample characteristics (age range, any condition or diagnosis, source [e.g., clinic-based, household-based, school-based, other])<br>• Study design<br>• Number of cycles per participant<br>• Number of cycles total<br>• Date of data collection<br>• Electronic data collection<br>• Validation methodology |
| 3. Instrument details | • Measure evaluated or validated as the primary measure<br>• Type of tool (full questionnaire, subscale, 1-2 questions, laboratory assay)<br>• Who fills out tool (patient at home, patient at clinic, clinician, researcher) |

| | |
|---|---|
| | • Measure or measures used as comparison group for primary measure under consideration<br>• All menstrual changes the instrument can measure<br>• Menstrual changes measurement validated in the study |
| 4. Measure quality criteria | • Conceptual and measurement model<br>• Reliability<br>• Content validity<br>• Construct validity<br>• Responsiveness/dynamism<br>• Sensitive nature of questions |
| 5. Clinical trial utility criteria | • Interpretability of results<br>• Transferability<br>• Participant burden<br>• Investigator burden |

For assessing measure quality and clinical trial utility, one author (SC) reviewed existing evaluation criteria and tools from the literature and guidance documents on selecting instruments for clinical trials (e.g., see Crossnohere *et al*., 2021 [10] for a recent overview) with input from the rest of the authors. After considering several alternatives (e.g., COSMIN Risk of Bias checklist [11], Francis *et al*.'s checklist to operationalize measurement characteristics of PRO measures [12], and the International Professional Society for Health Economics and Outcomes Research (ISPOR) PRO Good Research Practices Task Force guidance [13,14]), we determined these approaches did not meet our needs due to being too burdensome, too binary, or not specific to evaluation, respectively. We decided to follow the Patient-Reported Outcomes Tools: Engaging Users and Stakeholders (PROTEUS) Consortium recommendations to use International Society for Quality of Life Research (ISOQOL) standards for PRO measures [15,16]. We made two adjustments to the ISOQOL standards: (a) we added an attribute on sensitivity of questions given the topic of menstruation has a noted amount of stigma surrounding it [17]; and (b) we separated out participant burden from investigator burden given these two can differ greatly for instruments measuring menstrual changes. We categorized six attributes as related primarily to the quality of the instrument (i.e., **measure quality**: conceptual/measurement model, reliability, content validity, construct validity, responsiveness, and sensitive nature of questions) and four attributes as related primarily to the utility of the instrument in clinical trials (i.e., **clinical trial utility**: interpretability of results, the transferability of the instrument, participant burden, and investigator burden).

We scored each attribute of measure quality and clinical trial utility on a scale from 0 to 3, where 0 indicated there were **no data** reported on the attribute, 1 indicated **poor** measure quality/clinical trial utility of the attribute, 2 indicated **fair** measure quality/clinical trial utility of the attribute, and 3 indicated **good** measure quality/clinical trial utility of the attribute. Criteria for scoring of an attribute was defined in line with ISOQOL standards [16] and reviewed by measurement and clinical experts at FHI 360 and within the Global CIMC Task Force. We show the measure quality and clinical trial utility attributes and scoring criteria in Table D.

#### Table D: Measure quality and clinical trial utility scoring criteria*

| Attribute | Poor quality (1) | Fair quality (2) | Good quality (3) |
|---|---|---|---|
| **Measure quality** | | | |
| **Conceptual and Measurement Model**<br>Definition: The conceptual model provides a description and framework for targeted construct(s) in the measure. The measurement model maps individual measure items to the construct(s).<br>*Score 0 if not assessed in article.* | Minimal discussion of conceptual model or measurement model that maps measure items to the construct(s).<br>Or minimal discussion of intended population or context for measure use. | Some discussion of conceptual and/or measurement model that maps measure items to the construct(s).<br>Or some discussion of intended population and/or context for measure use. | Clearly defines and describes concept(s) included in model and intended population(s) and context for measure use.<br>Or clearly describes how concept(s) are organized into measurement model, including evidence for dimensionality of the measure, how items relate to each measured concept, and the relationship among concepts. |
| **Reliability**<br>Definition: The degree to which a measure is free from measurement error.<br>*Score 0 if not assessed in article.* | There is minimal evidence for measure reliability (e.g., internal consistency reliability, test-retest reliability, or item response theory) | Unclear or unjustified methodology used for assessing reliability.<br>Or, if used, reliability Cronbach α <0.70 for group-level comparisons without justification. | Methodology for collecting data is justified (e.g., a multi-item measure is assessed for internal consistency reliability and a single-item measure is assessed by test-retest reliability or item response theory).<br>Or, if used, reliability Cronbach α ≥0.70 for group-level comparisons. If lower, there is clear and appropriate justification. |
| **Content Validity**<br>Definition: The extent to which the measure includes the most relevant and important aspects | Minimal evidence participants or experts consider the measure relevant and comprehensive. | Some evidence participants and experts consider the measure relevant and/or comprehensive | Clear evidence participants and experts consider the measure relevant and comprehensive for the |

| Attribute | Poor quality (1) | Fair quality (2) | Good quality (3) |
|---|---|---|---|
| of a concept in the context of a given measurement application.<br>*Score 0 if not assessed in article.* | Or minimal documentation of methodology for evaluating content validity. | for the concept, population, and/or intended application.<br>Or some evidence of methodology used to evaluate content validity.<br>Or the paper mentions past validation research (i.e., focus groups, pilot studies, formative research) but does not provide detail on these studies. | concept, population, and intended application.<br>And clear evidence of methodology used to evaluate content validity, including for assessing the relevance of measured concept(s), comparing validation study sample to the wider target population, and justification for recall period. |
| **Construct Validity**<br>Definition: The degree to which scores on the measure relate to other measures (e.g., patient-reported or clinical indicators) in a manner that is consistent with theoretically derived a priori hypotheses concerning the concepts being measured.<br>*Score 0 if not assessed in article.* | Minimal evidence supporting pre-determined hypotheses related to construct validity. | Some evidence supporting pre-determined hypotheses related to construct validity. | Clear evidence supporting pre-defined hypotheses on the expected associations among other measures similar or dissimilar to the studied measure. |
| **Responsiveness/dynamism**<br>Definition: The extent to which a measure can detect changes in the construct being measured over time.<br>*Score 0 if not assessed in article.* | Minimal evidence the measure can detect changes consistent with pre-defined hypotheses related to responsiveness.<br>Or minimal evidence the measure can detect changes within or among participant groups. | Some evidence the measure can detect changes consistent with pre-defined hypotheses related to responsiveness.<br>Or some evidence the measure can detect changes within or among participant groups. | Clear evidence the measure can detect changes consistent with pre-defined hypotheses in the target population for the intended application.<br>And clear evidence the measure can detect changes within or among participant groups. |
| **Sensitive nature of items**<br>Definition: How measure addresses questions of sensitive topics, including those that are seen as intrusive, posing a threat of disclosure, or eliciting socially desirable answers.<br>*Score 0 if not assessed in article.* | Minimal evidence about measure or item sensitivity<br>Or evidence of sensitivity that may result in biased responses | Some evidence or discussion about measure or item sensitivity<br>Or some evidence of reduced sensitivity that would not result in biased responses | Clear evidence about measure or item sensitivity<br>And clear evidence of reduced sensitivity that would not result in biased responses |
| **Clinical trial utility** | | | |
| **Interpretability of results**<br>Definition: The degree to which one can easily understand a | Minimal evidence for interpreting results. | Some evidence for interpreting results. | Clear evidence of interpreting results, including |

| Attribute | Poor quality (1) | Fair quality (2) | Good quality (3) |
|---|---|---|---|
| measure's results (e.g., scores, levels).<br>*Score 0 if not provided in article.* | Or minimal evidence results are understood by relevant stakeholders. There is no clinically relevant minimum change or no assessment of clinical relevance. | Or some evidence results are understood by relevant stakeholders, including patients, clinicians, and/or researchers. There is an agreement on clinically relevant minimum change and/or assessment of clinical relevance. | differentiating between differing outcomes (e.g., high and low scores), and/or what constitutes a large or small change in the measured concept. And evidence results are clearly understood by multiple relevant stakeholders, including patients, clinicians, and researchers. There is an accepted clinically relevant minimum change. |
| **Transferability**<br>Definition: The degree to which the measure can be transferred between linguistic and sociocultural groups.<br>*Score 0 if not provided in article.* | Minimal evidence measurement properties are maintained across linguistic and/or cultural groups. | Some evidence measurement properties are maintained across linguistic and/or cultural groups. | Clear evidence measurement properties are maintained across linguistic or cultural groups, including qualitative testing of the translated measure. |
| **Participant Burden**<br>Definition: The time, effort, resource (e.g., use or ownership of smart phone, internet access refrigeration), and other demands placed on those to whom the measure is administered.<br>*Score 0 if not provided in article.* | Measure requires more than 20 minutes† to complete (>40 questions), requires data collection daily or multiple times a day, and/or multiple clinic visits or daily data collection outside the home. Or there is no information on expected participant time burden.<br>Or the measure requires resources not available to most participants.<br>Or there is minimal information on literacy demand of measure items or appropriateness for proposed context. | Measure requires between 15-20 minutes† to complete (20-40 questions), and/or one or two clinic visits, including those that are a burden to participant. Or there is limited information on expected participant time burden, including limited or no input from participant review panels. Or the measure may require some resources can be a barrier to some participants.<br>Or literacy demand of measure items is above a 6th grade level (i.e., >12-year-old) and not appropriately justified for proposed context. | Measure requires less than 15 minutes† to complete (<20 questions), no daily data collection, and no more than one clinic visit. Or there is an accurate description of the expected participant time burden with approval from participant review panels.<br>Or there are no resource barriers to participants.<br>And literacy demand of measure items is at a 6th grade level or lower (i.e., ≤12-year-old), or literacy level is appropriately justified for proposed context. |

| Attribute | Poor quality (1) | Fair quality (2) | Good quality (3) |
|---|---|---|---|
| **Investigator Burden**<br>Definition: The time, effort, resource, and other demands placed on those who administer the measure.<br>*Score 0 if not provided in article*. | There is a high burden on the data collection team due to: (a) data collector training being time or cost prohibitive with a lack of available training materials; (b) a high data monitoring burden to maintain quality data; (c) measure scoring being complex; or (d) measure inflexible or resource intensiveness (e.g., can only be interviewer-administered or requires tablet or computer).<br>Or there is minimal information on investigator burden. | There is a modest burden on the data collection team due to: (a) the time and cost of data collector training or lack of training materials; (b) data monitoring burden; (c) modest measure scoring complexity; or (d) the measure being either flexible or not resource intensive.<br>Or there is limited information on investigator burden. | There is a low burden on a data collection team due to (a) minimal requirement for data collector training and availability of training materials; (b) low data monitoring burden, (c) measure scoring being simple, or (d) the measure being flexible and not resource intensive (e.g., either measure is completed by the participant or is easily explained and completed).<br>Or there is an accurate description of the expected investigator burden. |

* Attributes and definitions from Reeve *et al*. 2013 [16] per PROTEUS-Trials Consortium guidance [15], with modified as specified in the text.
† Crossnohere *et al.*, 2021 [10].

#### Process for title/abstract screening, full text review, and data extraction

The authors met with the FHI 360 health sciences library team for a month to finalize the search strategy and then began weekly author meetings to discuss progress, questions, and discordance, and to document decisions and progress in a shared Word document. We began title/abstract screening with an 'inter-reviewer reliability' meeting where all authors completed title/abstract screening on the same 50 articles to establish and confirm group standards. Then, two authors independently screened each remaining title/abstract and two authors independently reviewed each relevant full text in Covidence. We resolved any discordance during weekly meetings via consensus conversations. We used the text translation feature of Google Translate to review abstracts not in English during screening, and we used the document translation feature of Google Translate and/or consulted a fluent colleague to review full text articles that were not in English. We used the notes and tag features in Covidence to document questions between meetings, consensus decisions during meetings, and any translation from Google Translate. We used Excel worksheets for data extraction. For instruments reported in more than one article, we concurrently extracted all articles on each instrument. We conducted data extraction with a fluent colleague for full text articles not in English. During title/abstract screening, full text review, and data extraction, when the authors had finished with approximately 5% of the articles, the following weekly author meeting included a specific discussion on the need for any clarifications or minor modifications to our inclusion/exclusion criteria for screenings/review or data extraction forms. After these '5% discussions', we made only minor clarifications to the inclusion/exclusion criteria and added or revised only a few fields in the data extraction forms.

#### Data analysis

Two authors (EH and SC) developed the initial analysis plan with input from the rest of the authors, and one author (EH) compiled all extracted data and conducted initial analyses with data checks by the rest of the authors. After data compilation, all authors conducted parts of the analysis. All analysis was conducted in Excel and included counts and frequencies, as well as specific analyses to assess (a) measure quality and (b) clinical trial utility. For these two outcomes, two authors (EH and SC) developed a scoring system with input from other authors in order to assign each instrument a measure quality score, a clinical trial utility score, and a total evidence score. For **measure quality scores** and **clinical trial utility scores**, we used an average of the highest score for each attribute of measure quality or clinical trial utility across all articles on an instrument. Because instruments could have more than one article providing data on measure quality and/or clinical trial utility and not every article evaluated all attributes, we did not include scores of zero (i.e., no data reported) in the measure quality and clinical trial utility scores. To reflect these differences in the number of articles and attributes reported in the article(s), we also calculated a total **evidence score**, which was the total of all scores—including zeros—across all attributes of measure quality and clinical trial utility. The total evidence scores, therefore, 'penalize' instruments for a lower level of evidence due to fewer articles or less attribute data and vice versa.

These three scores—measure quality (ranging from 1-3), clinical trial utility (ranging from 1-3), and total evidence (ranging 0+)—reflect different dimensions of an instrument. For example, two instruments might both have a score of 2.5 for measure quality, but one instrument might have an evidence score of 10 and the other, 100, indicating the latter has considerably more evidence and likely more certainty in the measure quality score. Alternately, two instruments may have similar measure quality and evidence scores, but one may have a clinical trial utility score of 1 and the other a score of 3, indicating the latter is likely better suited for use in clinical trials despite the similar levels of measure quality and evidence.

14. Patrick DL, Burke LB, Gwaltney CJ, Leidy NK, Martin ML, Molsen E, et al. Content validity--establishing and reporting the evidence in newly developed patient-reported outcomes (PRO)

instruments for medical product evaluation: ISPOR PRO Good Research Practices Task Force report: part 2--assessing respondent understanding. Value Health. 2011;14: 978–88. doi:10.1016/j.jval.2011.06.013

15. Snyder C, Crossnohere N, King M, Reeve BB, Bottomley A, Calvert M, et al. The PROTEUS-Trials Consortium: Optimizing the use of patient-reported outcomes in clinical trials. Clin Trials. 2022;19: 277–284. doi:10.1177/17407745221077691

16. Reeve BB, Wyrwich KW, Wu AW, Velikova G, Terwee CB, Snyder CF, et al. ISOQOL recommends minimum standards for patient-reported outcome measures used in patient-centered outcomes and comparative effectiveness research. Qual Life Res. 2013;22: 1889–905. doi:10.1007/s11136-012-0344-y

17. Johnston-Robledo I, Chrisler JC. The Menstrual Mark: Menstruation as Social Stigma. In: Bobel C, Winkler I, Fahs B, Hasson K, Kissling E, Roberts T, editors. The Palgrave Handbook of Critical Menstruation Studies. Singapore: Springer Singapore; 2020. pp. 181–199. doi:10.1007/978-981-15-0614-7_17

#### S3 Table. All articles included after title/abstract screening and full text review.

| Measures being evaluated/validated | First author | Year | Region | Country | Study Design | Ref. |
|---|---|---|---|---|---|---|
| **Studies validating/evaluating full instruments (n=133)** | | | | | | |
| Aberdeen Menorrhagia Severity Scale | Ruta | 1995 | Europe | United Kingdom | Cross-Sectional | [1] |
| Aberdeen Menorrhagia Severity Scale | Abu-Rafea | 2012 | Middle East | Saudi Arabia | Cross-Sectional | [2] |
| Adolescent Dysmenorrhic Self-Care Scale | Ching-Hsing | 2004 | Asia | Taiwan | Cross-Sectional | [3] |
| Adolescent Dysmenorrhic Self-Care Scale | Wong | 2013 | Asia | Hong Kong | Cross-Sectional | [4] |
| Alkaline Hematin Assay | van Eijkeren | 1986 | Europe | Netherlands | Prospective Cohort | [5] |
| Average cycle length - self report | Small | 2007 | North America | United States | Prospective Cohort | [6] |
| Bleeding and Pelvic Discomfort Scale | Hudgens | 2022 | Multiple | Multiple | Randomized Controlled Trial | [7] |
| Daily diary, menopause classification | Paramsothy | 2013 | North America | United States | Prospective Cohort | [8] |
| Daily diary, menstrual cycle length | Johannes | 2000 | North America | United States | Prospective Cohort | [9] |
| Dysmenorrhea Daily Diary | Nguyen | 2015 | North America | United States | Cross-Sectional | [10] |
| Dysmenorrhea Daily Diary | Nguyen | 2017 | Multiple | Multiple | Cross-Sectional | [11] |
| Dysmenorrhea Symptom Interference Scale | Chen | 2021 | North America | United States | Mixed methods | [12] |
| electronic Personal Assessment Questionnaire - Menstrual, Pain, and Hormonal | Gray | 2019 | Europe | United Kingdom | Cross-Sectional | [13] |
| Endometriosis Daily Diary | Guan | 2022 | North America | United States | Prospective Cohort | [14] |
| Endometriosis Daily Pain Impact Diary | Wyrwich | 2018 | North America | United States | Prospective Cohort | [15] |
| Endometriosis Health Profile-30 | Jones | 2001 | Europe | United Kingdom | Cross-Sectional | [16] |

| Measures being evaluated/validated | First author | Year | Region | Country | Study Design | Ref. |
|---|---|---|---|---|---|---|
| Endometriosis Health Profile-30 | Mengarda | 2008 | South America | Brazil | Cross-Sectional | [17] |
| Endometriosis Health Profile-30 | Grundstrom | 2020 | Europe | Sweden | Cross-Sectional | [18] |
| Endometriosis Health Profile-30 | Grundstrom | 2020 | Europe | Sweden | Cross-Sectional | [19] |
| Endometriosis Health Profile-30 | Jones | 2004 | Europe | United Kingdom | Prospective Cohort | [20] |
| Endometriosis Health Profile-30 | Jenkinson | 2008 | North America | United States | Cross-Sectional | [21] |
| Endometriosis Health Profile-30 | Hansen | 2022 | Europe | Denmark | Cross-Sectional | [22] |
| Endometriosis Health Profile-30 | Khong | 2010 | Oceania | Australia | Cross-Sectional | [23] |
| Endometriosis Health Profile-30 | Verket | 2018 | Europe | Norway | Cross-Sectional | [24] |
| Endometriosis Health Profile-30 | Jones | 2006 | Europe | United Kingdom | Cross-Sectional | [25] |
| Endometriosis Health Profile-30 | van de Burgt | 2011 | Europe | Netherlands | Case Control | [26] |
| Endometriosis Health Profile-30 | Van de Burgt | 2013 | Europe | Netherlands | Prospective Cohort | [27] |
| Endometriosis Health Profile-30 | Wickstrom | 2017 | Europe | Sweden | Prospective Cohort | [28] |
| Endometriosis Health Profile-30 | Nojomi | 2011 | Middle East | Iran | Cross-Sectional | [29] |
| Endometriosis Health Profile-30 | Mansor | 2023 | Asia | Malaysia | Cross-Sectional | [30] |
| Endometriosis Health Profile-30 | Maiorana | 2012 | Europe | Italy | Cross-Sectional | [31] |
| Endometriosis Health Profile-30 | Marí-Alexandre | 2022 | Europe | Spain | Cross-Sectional | [32] |
| Endometriosis Health Profile-30 | Nogueria-Silva | 2015 | Europe | Portugal | Cross-Sectional | [33] |
| Endometriosis Health Profile-30 | Chauvet | 2017 | Europe | France | Cross-Sectional | [34] |
| Endometriosis Health Profile-30 | Jia | 2013 | Asia | China | Cross-Sectional | [35] |

| Measures being evaluated/validated | First author | Year | Region | Country | Study Design | Ref. |
|---|---|---|---|---|---|---|
| Endometriosis Health Profile-5 | Jones | 2004 | Europe | United Kingdom | Cross-Sectional | [36] |
| Endometriosis Health Profile-5 | Fauconnier | 2017 | Europe | France | Case Control | [37] |
| Endometriosis Health Profile-5 | Aubry | 2017 | Europe | France | Prospective Cohort | [38] |
| Endometriosis Health Profile-5 | Mikuš | 2023 | Europe | Croatia | Prospective Cohort | [39] |
| Endometriosis Health Profile-5 | Selcuk | 2015 | Europe | Turkey | Case Control | [40] |
| Endometriosis Impact Questionnaire | Moradi | 2019 | Oceania | Australia | Cross-Sectional | [41] |
| Endometriosis Impact Scale | Gater | 2020 | Multiple | Multiple | Mixed methods | [42] |
| Endometriosis Pain and Bleeding Diary | Deal | 2010 | North America | United States | Prospective Cohort | [43] |
| Endometriosis Pain Daily Diary | Van Nooten | 2018 | Multiple | Multiple | Mixed methods | [44] |
| Endometriosis Reproductive Health Questionnaire | Namazi | 2021 | Middle East | Iran | Mixed methods | [45] |
| Endometriosis Self-Assessment Tool | Cho | 2022 | Asia | South Korea | Cross-Sectional | [46] |
| Endometriosis Treatment Satisfaction Questionnaire | Deal | 2010 | North America | United States | Randomized Controlled Trial | [47] |
| ENDOPAIN-4D | Fauconnier | 2018 | Europe | France | Mixed methods | [48] |
| ENDOPAIN-4D | Puchar | 2021 | Europe | France | Prospective Cohort | [49] |
| ENDOPAIN-4D | Ahmadpour | 2022 | Middle East | Iran | Cross-Sectional | [50] |
| EndoWheel | As-Sanie | 2021 | North America | United States | Mixed methods | [51] |
| Fibroid Symptom Diary | Deal | 2011 | North America | United States | Mixed methods | [52] |
| Functional and Emotional Measure of Dysmenorrhea | Li | 2012 | Asia | China | Cross-Sectional | [53] |

| Measures being evaluated/validated | First author | Year | Region | Country | Study Design | Ref. |
|---|---|---|---|---|---|---|
| Injustice Experience Questionnaire-Chronic and the Contribution of Perceived Injustice | Yamada | 2019 | Asia | Japan | Prospective Cohort | [54] |
| Mansfield-Voda-Jorgensen Menstrual Bleeding Scale | Mansfield | 2004 | North America | United States | Prospective Cohort | [55] |
| Measure compilation (Olliges) | Olliges | 2021 | Europe | Germany | Case Control | [56] |
| Menorrhagia Impact Questionnaire | Bushnell | 2010 | North America | United States | Case Control | [57] |
| (Menorrhagia) Multi-Attribute Utility Score | Shaw | 1998 | Europe | United Kingdom | Mixed methods | [58] |
| (Menorrhagia) Multi-Attribute Utility Score | Habiba | 2010 | Europe | United Kingdom | Prospective Cohort | [59] |
| (Menorrhagia) Multi-Attribute Utility Score | Pattison | 2011 | Europe | United Kingdom | Randomized Controlled Trial | [60] |
| Menstrual Attitudes Questionnaire | Brooks-Gunn | 1980 | North America | United States | Cross-Sectional | [61] |
| Menstrual Attitudes Questionnaire | Bramwell | 2002 | Europe; Asia | United Kingdom; India | Cross-Sectional | [62] |
| Menstrual Attitudes Questionnaire | Firat | 2009 | Europe; Asia | Turkey | Cross-Sectional | [63] |
| Menstrual Attitudes Questionnaire | Bargiota | 2016 | Europe | Greece | Cross-Sectional | [64] |
| Menstrual Attitudes Questionnaire | Kawata | 2022 | Asia | Nepal | Cross-Sectional | [65] |
| Menstrual Bleeding Questionnaire | Rezende | 2023 | South America | Brazil | Prospective Cohort | [66] |
| Menstrual Bleeding Questionnaire | Matteson | 2015 | North America | United States | Prospective Cohort; Cross-Sectional | [67] |
| Menstrual Bleeding Questionnaire | Pike | 2021 | North America | Canada | Cross-Sectional, Prospective Cohort | [68] |
| Menstrual Bleeding Questionnaire | Rodpetch | 2021 | Asia | Thailand | Cross-Sectional | [69] |
| Menstrual Blood Loss Score Questionnaire | Toxqui | 2014 | Europe | Spain | Prospective Cohort | [70] |

| Measures being evaluated/validated | First author | Year | Region | Country | Study Design | Ref. |
|---|---|---|---|---|---|---|
| Menstrual Collection | Chimbira | 1980 | Europe | United Kingdom | Prospective Cohort | [71] |
| Menstrual Collection | Gleeson | 1993 | Europe | Ireland | Prospective Cohort | [72] |
| Menstrual Collection | Gannon | 1996 | Europe | United Kingdom | Prospective Cohort | [73] |
| Menstrual Collection | Fraser | 2001 | Oceania | Australia | Prospective Cohort | [74] |
| Menstrual Collection | Gudmundsdottir | 2009 | Europe | Iceland | Prospective Cohort | [75] |
| Menstrual Distress Questionnaire (Moos) | Moos | 1968 | North America | United States | Cross-Sectional | [76] |
| Menstrual Distress Questionnaire (Moos) | Lee | 2009 | Asia | China | Cross-Sectional | [77] |
| Menstrual Distress Questionnaire (Vannuccini) | Vannuccini | 2021 | Europe | Italy | Cross-Sectional | [78] |
| Menstrual Distress Questionnaire (Vannuccini) | Cassioli | 2023 | Europe; Oceania | Multiple | Cross-Sectional | [79] |
| Menstrual Health Instrument | Shin | 2018 | Asia | South Korea | Mixed methods | [80] |
| Menstrual Health Seeking Behaviors Questionnaire | Darabi | 2018 | Middle East | Iran | Cross-Sectional | [81] |
| Menstrual Hygiene Management Scale | Ramaiya | 2020 | Asia | India | Mixed methods | [82] |
| Menstrual Insecurity Tool | Caruso | 2020 | Asia | India | Mixed methods | [83] |
| Menstrual Joy Questionnaire | Aubeeluck | 2002 | Europe | United Kingdom | Cross-Sectional | [84] |
| Menstrual Practices Questionnaire | Hennegan | 2020 | Africa | Uganda | Cross-Sectional | [85] |
| Menstrual Record and Recall | Heath | 1999 | Oceania | New Zealand | Cross-Sectional | [86] |
| Menstrual Self-Evaluation Scale | Roberts | 2004 | North America | United States | Cross-Sectional | [87] |
| Menstruation-Related Activity Restrictions Questionnaire | Garg | 2021 | Asia | India | Cross-Sectional | [88] |

| Measures being evaluated/validated | First author | Year | Region | Country | Study Design | Ref. |
|---|---|---|---|---|---|---|
| Military Women's Attitudes Toward Menstrual Suppression Scale | Trego | 2009 | North America | United States | Mixed methods | [89] |
| New Zealand Survey of Adolescent Girls' Menstruation | Farquhar | 2009 | Oceania | New Zealand | Cross-Sectional | [90] |
| Numeric Rating Scale | deArruda | 2022 | South America | Brazil | Prospective Cohort | [91] |
| Numeric Rating Scale | Pokrzywinski | 2021 | North America | Multiple | Randomized Controlled Trial | [92] |
| Pain Drawing | Rodrigues | 2022 | South America | Brazil | Cross-Sectional | [93] |
| Period ImPact and Pain Assessment | Parker | 2022 | Oceania | Australia | Cross-Sectional | [94] |
| PERIOD-QOL | Lancastle | 2023 | Europe | United Kingdom | Cross-Sectional | [95] |
| Pictorial Blood Loss Assessment Charts & Menstrual Pictograms | Higham | 1990 | Europe | United Kingdom | Cross-Sectional | [96] |
| Pictorial Blood Loss Assessment Charts & Menstrual Pictograms | Janssen | 1995 | Europe | Netherlands | Cross-Sectional | [97] |
| Pictorial Blood Loss Assessment Charts & Menstrual Pictograms | Barr | 1999 | Africa | Nigeria | Cross-Sectional | [98] |
| Pictorial Blood Loss Assessment Charts & Menstrual Pictograms | Reid | 2000 | Europe | United Kingdom | Prospective Cohort | [99] |
| Pictorial Blood Loss Assessment Charts & Menstrual Pictograms | Wyatt | 2001 | Europe | United Kingdom | Prospective Cohort | [100] |
| Pictorial Blood Loss Assessment Charts & Menstrual Pictograms | Sanchez | 2012 | North America | United States | Prospective Cohort | [101] |
| Pictorial Blood Loss Assessment Charts & Menstrual Pictograms | Larsen | 2013 | North America | United States | Randomized Controlled Trial | [102] |
| Pictorial Blood Loss Assessment Charts & Menstrual Pictograms | Magnay | 2013 | Europe | United Kingdom | Laboratory | [103] |
| Pictorial Blood Loss Assessment Charts & Menstrual Pictograms | Magnay | 2014 | Europe | United Kingdom | Prospective Cohort | [104] |
| Pictorial Blood Loss Assessment Charts & Menstrual Pictograms | Hald | 2014 | Europe | Norway | Retrospective Cohort | [105] |

| Measures being evaluated/validated | First author | Year | Region | Country | Study Design | Ref. |
|---|---|---|---|---|---|---|
| Pictorial Blood Loss Assessment Charts & Menstrual Pictograms and Uterine Fibroid Daily Bleeding Diary | Haberland | 2020 | Europe | Germany | Randomized Controlled Trial | [106] |
| Prospective self report, menstrual regularity | Weller | 1998 | Middle East | Israel | Prospective Cohort | [107] |
| Quantitative model for menstrual blood loss | Schumacher | 2012 | Multiple | Multiple | Retrospective Cohort | [108] |
| Retrospective self-report, last menstrual period | Wegienka | 2005 | North America | United States | Prospective Cohort | [109] |
| Retrospective self report, menstrual discomfort | Jukic | 2008 | North America | United States | Prospective Cohort | [110] |
| Retrospective self report, menstrual length (Bachand) | Bachand | 2009 | North America | United States | Prospective Cohort | [111] |
| Retrospective self report, menstrual length (Small & Jukic) | Jukic | 2007 | North America | United States | Prospective Cohort | [112] |
| SAMANTA Questionnaire | Calaf | 2020 | Europe | Spain | Cross-Sectional | [113] |
| SAMANTA Questionnaire | Perelló-Capo | 2023 | Europe | Spain | Prospective Cohort | [114] |
| Spanish Society of Contraception Quality-of-Life | Pérez-Campos | 2011 | Europe | Spain | Prospective Cohort | [115] |
| Squeezing Pain Bulb | Kantarovich | 2021 | North America | United States | Cross-Sectional | [116] |
| Uterine Fibroid Symptom and Quality of Life Questionnaire | Spies | 2002 | North America | United States | Cross-Sectional | [117] |
| Uterine Fibroid Symptom and Quality of Life Questionnaire | Harding | 2008 | North America | United States | Prospective Cohort | [118] |
| Uterine Fibroid Symptom and Quality of Life Questionnaire | Silva | 2016 | South America | Brazil | Case Control | [119] |
| Uterine Fibroid Symptom and Quality of Life Questionnaire | Oliveira Brito | 2017 | South America | Brazil | Cross-Sectional | [120] |
| Uterine Fibroid Symptom and Quality of Life Questionnaire | Coyne | 2017 | North America | United States | Randomized Controlled Trial | [121] |
| Uterine Fibroid Symptom and Quality of Life Questionnaire | Coyne | 2019 | North America | Multiple | Randomized Controlled Trial | [122] |
| Uterine Fibroid Symptom and Quality of Life Questionnaire | Yeung | 2019 | Asia | Hong Kong | Cross-Sectional | [123] |

| Measures being evaluated/validated | First author | Year | Region | Country | Study Design | Ref. |
|---|---|---|---|---|---|---|
| Uterine Fibroid Symptom and Quality of Life Questionnaire | Calaf | 2020 | Europe | Spain | Cross-Sectional | [124] |
| Uterine Fibroid Symptom and Quality of Life Questionnaire | Keizer | 2021 | Europe | Netherlands | Cross-Sectional | [125] |
| Visual Analogue Scales: Pain | Gerlinger | 2012 | Europe | Germany | Retrospective Cohort | [126] |
| Visual Analogue Scales: Pain | Gerlinger | 2010 | Europe | Multiple | Retrospective Cohort | [127] |
| Visual Analogue Scales: Bleeding | Perelló | 2022 | Europe | Spain | Retrospective Cohort | [128] |
| World Health Organization Disability Assessment Schedule 2.0 | deArruda | 2023 | South America | Brazil | Cross-Sectional | [129] |
| Working Ability, Location, Intensity, Days of Pain, Dysmenorrhea Score | Teherán | 2018 | South America | Colombia | Cross-Sectional | [130] |
| Working Stressors and Coping Strategies Associated with Menstrual Symptoms Among Nurses | Yu | 2021 | Asia | Taiwan | Cross-Sectional | [131] |
| World Endometriosis Research Foundation Endometriosis Phenome and Biobanking Harmonisation Project Standard Questionnaire | Vitonis | 2014 | Multiple | Multiple | Other | [132] |
| World Endometriosis Research Foundation Endometriosis Phenome and Biobanking Harmonisation Project Standard Questionnaire | Dimentberg | 2021 | North America | Canada | Prospective Cohort | [133] |
| **Studies validating/evaluating applicable subscales (n=19)** | | | | | | |
| Low Energy Availability in Females Questionnaire | Melin | 2014 | Europe | Multiple | Prospective Cohort | [134] |
| Low Energy Availability in Females Questionnaire | deMaria | 2021 | South America | Brazil | Prospective Cohort | [135] |
| Menstrual Cycle-Related Signs and Symptoms Questionnaire | Sutthibut | 2021 | Asia | Thailand | Cross-Sectional | [136] |
| Menstrual Symptom Questionnaire | Chesney | 1975 | North America | United States | Cross-Sectional | [137] |
| Menstrual Symptom Questionnaire | Nelson | 1984 | North America | United States | Cross-Sectional | [138] |
| Menstrual Symptom Questionnaire | Negriff | 2009 | North America | United States | Cross-Sectional | [139] |

| Measures being evaluated/validated | First author | Year | Region | Country | Study Design | Ref. |
|---|---|---|---|---|---|---|
| Midlife Women's Symptom Index | Im | 2006 | North America | United States | Cross-Sectional | [140] |
| ORTHO Birth Control Satisfaction Assessment Tool | Mathias | 2006 | North America | United States | Mixed methods | [141] |
| ORTHO Birth Control Satisfaction Assessment Tool | Colwell | 2006 | North America | United States | Cross-Sectional | [142] |
| Ovulation and Menstruation Health Questionnaire | Mahalingaiah | 2020 | North America | United States | Mixed methods | [143] |
| Polycystic Ovary Syndrome Quality of Life Scale | Saei Ghare Naz | 2023 | Middle East | Iran | Mixed methods | [144] |
| Polycystic Ovary Syndrome Quality of Life Scale | Cronin | 1998 | North America | United States | Mixed methods | [145] |
| Polycystic Ovary Syndrome Quality of Life Scale | Jones | 2004 | Europe | United Kingdom | Cross-Sectional | [146] |
| Polycystic Ovary Syndrome Quality of Life Scale | Guyatt | 2004 | Europe | United Kingdom | Randomized Controlled Trial | [147] |
| Polycystic Ovary Syndrome Quality of Life Scale | Jedel | 2008 | Europe | Sweden | Randomized Controlled Trial | [148] |
| Polycystic Ovary Syndrome Quality of Life Scale | Bazarganipour | 2012 | Middle East | Iran | Cross-Sectional | [149] |
| Polycystic Ovary Syndrome Quality of Life Scale | Bazarganipour | 2013 | Middle East | Iran | Cross-Sectional | [150] |
| Polycystic Ovary Syndrome Quality of Life Scale | Chung | 2016 | Asia | China | Cross-Sectional | [151] |
| Women Shift Workers Reproductive Health Questionnaire | Nikpour | 2020 | Middle East | Iran | Mixed methods | [152] |
| **Studies validating/evaluating instruments with applicable items/questions (n=14)** | | | | | | |
| Adolescent Menstrual Attitude Questionnaire | Morse | 1993 | North America | Canada | Not assessed | [153] |
| Adolescent Menstrual Attitude Questionnaire | Morse | 1993 | North America | Canada | Not assessed | [154] |
| CARDIA Women's Reproductive Health Questionnaire | Whitham | 2013 | North America | United States | Not assessed | [155] |

| Measures being evaluated/validated | First author | Year | Region | Country | Study Design | Ref. |
|---|---|---|---|---|---|---|
| Clinical Tool for Diagnosis of PCOS | Pedersen | 2007 | North America | Canada | Not assessed | [156] |
| Clinically Validated Scores for Endometriosis Diagnosis | Chapron | 2022 | Europe | France | Not assessed | [157] |
| Experience Sampling Method | van Barneveld | 2023 | Europe | Netherlands | Not assessed | [158] |
| Immune Thrombocytopenic Purpura Patient Assessment Questionnaire | Mathias | 2007 | Multiple | Multiple | Not assessed | [159] |
| Painful Periods Screening Tool | DiBenedetti | 2018 | North America | United States | Not assessed | [160] |
| Polycystic Ovary Syndrome Questionnaire-50 | Nasiri-Amiri | 2016 | Middle East | Iran | Not assessed | [161] |
| Polycystic Ovary Syndrome Questionnaire-50 | Stevanovic | 2019 | Europe | Serbia | Not assessed | [162] |
| Self-Efficacy in Addressing Menstrual Needs Scale | Hunter | 2022 | Asia | Bangladesh | Not assessed | [163] |
| Stellenbosch Endometriosis Quality of Life Measure | Rizwana | 2018 | Africa | South Africa | Not assessed | [164] |
| Structured Endometriosis Questionnaire | Hackethal | 2011 | Europe | Germany | Not assessed | [165] |
| The International Spinal Cord Injury Female Sexual and Reproductive Function | Alexander | 2011 | Multiple | Multiple | Not assessed | [166] |
| **Studies validating/evaluating general instruments in menstruating populations (n=8)** | | | | | | |
| Health Related Productivity Questionnaire | Pokrzywinski | 2020 | Multiple | Multiple | Not assessed | [167] |
| Patient-Generated Index of Quality of Life | Ruta | 1999 | Europe | United Kingdom | Not assessed | [168] |
| Patient-Reported Outcomes Measurement Information System | Schneider | 2013 | North America | United States | Not assessed | [169] |
| Patient-Reported Outcomes Measurement Information System | Pokrzywinski | 2020 | North America | Multiple | Not assessed | [170] |
| Survey of Pain Attitudes | Ferreira-Valente | 2019 | Europe | Portugal | Not assessed | [171] |
| Women's Health Questionnaire | Hunter | 1992 | Europe | United Kingdom | Not assessed | [172] |

| Measures being evaluated/validated | First author | Year | Region | Country | Study Design | Ref. |
|---|---|---|---|---|---|---|
| Women's Health Questionnaire | Hunter | 2000 | Europe | United Kingdom | Not assessed | [173] |
| Women's Health Questionnaire | Colantonio | 2011 | North America | Canada | Not assessed | [174] |

70. Toxqui L, Pérez-Granados AM, Blanco-Rojo R, Wright I, Vaquero MP. A simple and feasible questionnaire to estimate menstrual blood loss: relationship with hematological and

gynecological parameters in young women. BMC Womens Health. 2014;14: 71. doi:10.1186/1472-6874-14-71

131. Yu S-C, Hsu H-P, Guo J-L, Chen S-F, Huang S-H, Chen Y-C, et al. Exploration of the experiences of working stressors and coping strategies associated with menstrual symptoms among nurses with

shifting schedules: a Q methodology investigation. BMC Nurs. 2021;20: 238. doi:10.1186/s12912-021-00759-0

#### S4 Table. Characteristics of sub-scales, items, and general instruments.

| Full Name of instrument | Available Languages | Available Electronically?* | BLEEDING | | | | BLOOD | | | UTERINE PAIN | PERCEP-TIONS | Subscale Topic or Wording of Questions | Ref |
|---|---|---|---|---|---|---|---|---|---|---|---|---|---|
| | | | Duration | Volume | Frequency | Regularity | Color | Consistency | Smell | | | | |
| **Instruments with subscales on menstrual changes (n=8)** | | | | | | | | | | | | | |
| Low Energy Availability in Females Questionnaire | Danish, English, Portuguese, Swedish | Yes | X | X | X | X | | | | | | Menstrual function and use of contraceptives | [134,135] |
| Menstrual Cycle-Related Signs and Symptoms Questionnaire | Thai | No | | X | | | X | X | X | | X | Menstrual cycle-related signs and symptoms (section 1) | [136] |
| Menstrual Symptom Questionnaire | English | No | | | | | | | | X | | Spasmodic dysmenorrhea, also called menstrual discomfort or factor 1: abdominal pain in later versions of the too | [137–139] |
| Midlife Women's Symptom Index | English | No | | X | X | X | | | | | | Self-reported menopausal status (includes last menstrual cycle, menstrual regularity, and menstrual flow) | [140] |
| ORTHO Birth Control Satisfaction Assessment Tool | English | No | X | X | X | | | | | X | X | Menstrual impact and lifestyle impact | [141,142] |
| Ovulation and Menstruation Health Survey | English | Yes | X | X | | X | | | | | | Menstrual cycle questions | [143] |
| Polycystic Ovary Syndrome Quality of Life Scale | Chinese, English, Persian, Swedish | No | | | | X | | | | X | | Menstrual problems (menstruation or menstrual) | [144–151] |
| Women Shift Workers Reproductive Health Questionnaire | Persian | No | | | | X | | | | X | | Menstruation | [152] |
| **Instruments with items or questions on menstrual changes (n=13)** | | | | | | | | | | | | | |
| Adolescent Menstrual Attitude Questionnaire | English | No | | | | X | | | | X | X | [strongly disagree; disagree; don't care, are not sure, or do not know; agree with the statement; strongly]: "I worry a lot about my periods starting unexpectedly."; "I do not feel any different than usual when I menstruate."; "I feel okay when I get my period."; "When I have my period, I feel good."; "When I get my period, I feel sick."; "Cramps during my period are very painful."; "I feel ugly and gross when I have my period."; "When I am menstruating I feel the same." | [153,154] |
| CARDIA Women's Reproductive Health Questionnaire | English | Yes | | | | X | | | | | | "During the past 12 months, have your menstrual cycles been regular at least half the time (excluding times when you were on birth control pills, pregnant, or nursing)?: [no, yes, not sure]?" | [155] |
| Clinical Tool for Diagnosis of PCOS | English | No | | | X | | | | | | | "Please answer this question NOT INCLUDING any time spent pregnant, receiving birth control pills or injections, after menopause, or after having both ovaries or the uterus surgically removed: Between the ages of 16 and 40, about how long was your average menstrual cycle (time from first day of one period to the first day of the next period)? Select ONE only: [<25 d, 25-34 d, 35-60 d, more than 60 d , totally variable]" | [156] |
| Clinically Validated Scores for Endometriosis Diagnosis** | French | No | | | | | | | | X | | Visual analogue scale (range: 0-10): dysmenorrhea ≥6 | [157] |
| Experience Sampling Method | Dutch | Yes | | | | | | | | X | | "I suffer from abdominal pain."; "I feel pain when I am standing/walking." | [158] |
| Immune Thrombocytopenic Purpura Patient Assessment Questionnaire (ITP-PAQ) | Multi-Site Study | No | X | X | | | | | | X | X | "Thinking about your last period…. How bothered were you by heavier bleeding before having ITP?"; "How bothered were you by bleeding for more days than before having ITP?"; "How bothered were you by more pain before ITP? " [5-point Likert scale from "extremely" to "not at all"] | [159] |
| Painful Periods Screening Tool | English | No | | | | | | | | X | X | "Do you often experience pelvic/abdominal or lower back pain before or during your periods that limits your activities or requires medication?"; Do you sometimes avoid sexual intercourse to avoid pain? [Yes, No, Not applicable, I am not sexually active]" | [160] |
| Polycystic Ovary Syndrome Questionnaire-50 | Persian, Serbian | No | | | | | | | | | X | "In the past 4 weeks have you ever...Felt concerned about menstruation at long intervals?" [Y/N] | [161,162] |
| Self-Efficacy in Addressing Menstrual Needs Scale | Bengali | Yes | X | | | | | | | X | X | "How confident are you that you can count/keep track of your period days?"; "How confident are you that you can usually reduce your abdominal pain by a small amount?"; "How confident are you that you can usually reduce most of your abdominal pain?"; "How confident are you that you can usually reduce your abdominal pain completely?" | [163] |

| Full Name of instrument | Available Languages | Available Electronically?* | BLEEDING | | | | BLOOD | | | UTERINE PAIN | PERCEP-TIONS | Subscale Topic or Wording of Questions | Ref |
|---|---|---|---|---|---|---|---|---|---|---|---|---|---|
| | | | Duration | Volume | Frequency | Regularity | Color | Consistency | Smell | | | | |
| Stellenbosch Endometriosis Quality of Life Measure | English | No | | | | | | X | | | X | "I was concerned about the clots in my period."; "I was worried that my period was not normal."; "I felt that my period drained me." [not applicable, not at all, a little bit, somewhat, quite a bit, very much] | [164] |
| Structured Endometriosis Questionnaire | English, German | Yes | | X | | X | | | | X | | "Do you have a regular menstrual cycle? [Y/N]"; "How long is your average period?"; "How long is your average bleeding?"; "When was the first day of your last period?"; "How do you estimate the intensity of the last menstrual bleeding?"; "Do you have pain in connection with you menstrual bleeding? [Y/N], If yes, when do you feel pain [previously, meanwhile, afterwards] How strong do you feel the pain on a scale from 0 to 10?" | [165] |
| The International Spinal Cord Injury Female Sexual and Reproductive Function | English | No | X | X | X | | | | | | | "How would you rate your current menstruation pattern? [Normal, Reduced/altered, Absent, Unknown, Not applicable]" | [166] |
| Uterine Fibroid Daily Bleeding Diary | Multi-Site Study | No | | X | | | | | | | | "Rate the severity of any vaginal bleeding in the past 24 hours. [No vaginal bleeding, Spotting, Mild, Moderate, Severe, Very severe]" | [106] |
| **General instruments validate in menstruating populations (n=5)** | | | | | | | | | | | | | |
| Health Related Productivity Questionnaire | Multi-Site Study | Yes | | | | | | | | | X | NA (General questions applied to menstruating population but no questions specific to menstruation) | [167] |
| Patient-Generated Index of Quality of Life | English | No | | | | | | | | | X | "We would like you to think of the most important areas of your life that are affected by your Menorrhagia. Please write up to FIVE areas in the boxes below." | [168] |
| Patient-Reported Outcomes Measurement Information System | English | Yes | X | | | | | | | X | X | NA (General questions applied to menstruating population but no questions specific to menstruation) | [169,170] |
| Survey of Pain Attitudes | Portuguese | Yes | | | | | | | | X | X | NA (General questions applied to menstruating population but no questions specific to menstruation) | [171] |
| Women's Health Questionnaire | English | No | | X | | | | | | X | | "Please indicate how you are feeling now, or how you have been feeling THE LAST FEW DAYS, by putting a tick in the correct box in the answer to each of the following items [Yes definitely, yes sometimes, no not much, no not at all]: 'I have heavy periods.'; 'My breasts feel tender or uncomfortable.'; 'I have abdominal cramps or discomfort.'" | [172–174] |

*According to publications, "Yes" indicates either fully or partly electronic

**All tools or subscales were designed to be completed by patients or participants, except for the Clinically Validated Scores for Endometriosis Diagnosis

70. Toxqui L, Pérez-Granados AM, Blanco-Rojo R, Wright I, Vaquero MP. A simple and feasible questionnaire to estimate menstrual blood loss: relationship with hematological and

gynecological parameters in young women. BMC Womens Health. 2014;14: 71. doi:10.1186/1472-6874-14-71

131. Yu S-C, Hsu H-P, Guo J-L, Chen S-F, Huang S-H, Chen Y-C, et al. Exploration of the experiences of working stressors and coping strategies associated with menstrual symptoms among nurses with

shifting schedules: a Q methodology investigation. BMC Nurs. 2021;20: 238. doi:10.1186/s12912-021-00759-0
